# The new normal? Dynamics and scale of the SARS-CoV-2 variant Omicron epidemic in England

**DOI:** 10.1101/2022.03.29.22273042

**Authors:** Oliver Eales, Leonardo de Oliveira Martins, Andrew J. Page, Haowei Wang, Barbara Bodinier, David Tang, David Haw, Jakob Jonnerby, Christina Atchison, Deborah Ashby, Wendy Barclay, Graham Taylor, Graham Cooke, Helen Ward, Ara Darzi, Steven Riley, Paul Elliott, Christl A. Donnelly, Marc Chadeau-Hyam

## Abstract

The SARS-CoV-2 pandemic has been characterised by the regular emergence of genomic variants which have led to substantial changes in the epidemiology of the virus. With natural and vaccine-induced population immunity at high levels, evolutionary pressure favours variants better able to evade SARS-CoV-2 neutralising antibodies. The Omicron variant was first detected in late November 2021 and exhibited a high degree of immune evasion, leading to increased infection rates in many countries. However, estimates of the magnitude of the Omicron wave have relied mainly on routine testing data, which are prone to several biases. Here we infer the dynamics of the Omicron wave in England using PCR testing and genomic sequencing obtained by the REal-time Assessment of Community Transmission-1 (REACT-1) study, a series of cross-sectional surveys testing random samples of the population of England. We estimate an initial peak in national Omicron prevalence of 6.89% (5.34%, 10.61%) during January 2022, followed by a resurgence in SARS-CoV-2 infections in England during February-March 2022 as the more transmissible Omicron sub-lineage, BA.2 replaced BA.1 and BA.1.1. Assuming the emergence of further distinct genomic variants, intermittent epidemics of similar magnitude as the Omicron wave may become the ‘new normal’.

## Introduction

Since late 2020 SARS-CoV-2 variants of concern (VOCs) have emerged regularly [1–4] leading to substantial changes in national, regional and global dynamics of the COVID-19 pandemic. On 24 November 2021 a new PANGO lineage [5] B.1.1.529 was designated, consisting of genomes sequenced in South Africa and Botswana in the prior week [6], and declared the Omicron VOC by the World Health Organization [7]. Though the Omicron variant has been found to cause less severe disease than previous variants [8,9], it has also been shown to exhibit a large number of mutations [10] including 15 in the receptor binding domain that has allowed it to escape a majority of pre-existing SARS-CoV-2 neutralising antibodies [11]. Rising incidence in South Africa, following Omicron’s emergence, revealed a greater rate of transmission relative to previously dominant VOCs [6]. This has been linked to immune evasion [6,12], including a reduction in the effectiveness of COVID-19 vaccines against Omicron infection [13] and an increased ability to reinfect previously-infected individuals [14]. The increased growth rate has been linked to both a shorter generation time [15] and a greater number of transmission events per generation [16]. Despite many countries imposing strict travel bans, Omicron rapidly disseminated worldwide, with confirmed cases in 171 countries by 20 January 2022 [17].

However, the magnitude of the Omicron wave is not apparent in most countries since testing captures an unknown proportion of infections and is prone to bias due to changing testing capacities and variable/differential test-seeking behaviour [18]. It is likely that in some countries high levels of Omicron infections have saturated testing capacity [19] introducing further bias into estimates of Omicron’s dynamics.

Representative community surveys can avoid such biases and accurately measure the prevalence of the virus, with fewer overall tests required [20]. Here, we use data from the REal-time Assessment of Community Transmission-1 (REACT-1) study that has tested randomly selected cross-sections of the population of England approximately monthly since May 2020 [21]. We use overall swab-positivity and genomic sequencing from rounds 14 to 18 (9 September 2021 to 1 March 2022) of REACT-1 to describe the dynamics of the Omicron wave in England as it replaced the previously dominant Delta variant. We further explore the diversity of Omicron sub-lineages in round 16 (23 November - 14 December), 17 (5 January - 20 January) and 18 (8 February - 1 March) and how they have contributed to the overall dynamics.

### Omicron Delta competition

Within the REACT-1 samples we estimated Omicron prevalence of 0.11% (0.07%, 0.16%) by 7 December 2021 (Fig. 1A), three weeks after the first confirmed Omicron case in England was sampled (16 November, linked to recent travel) [22]. At the same date Delta, which had been at a steady high prevalence for the preceding 3 months, was estimated to be approximately twelve-fold higher at 1.31% (1.17%, 1.47%). Though the Omicron variant was likely introduced to England by international travel from Southern Africa [22], we find greater levels of similarity between REACT-1 sequences and sequences sampled in the USA, Germany and France (Sup Fig. 1) with inferred high rates of importation/exportation from/to these countries (Sup Fig. 2). This likely reflects greater rates of transmission between England and USA/Europe after Omicron was globally disseminated. The proportion of cases in England linked to Omicron rapidly increased, reaching 50% by 14 December (13 December, 16 December) 2021 and 90% by 23 December (20 December, 26 December) 2021 (Fig. 1B). The last Delta sample in REACT-1 (up to round 18) was detected on 14 February 2022 when the proportion of cases linked to Omicron was greater than 99.82%.

**Figure 1:**
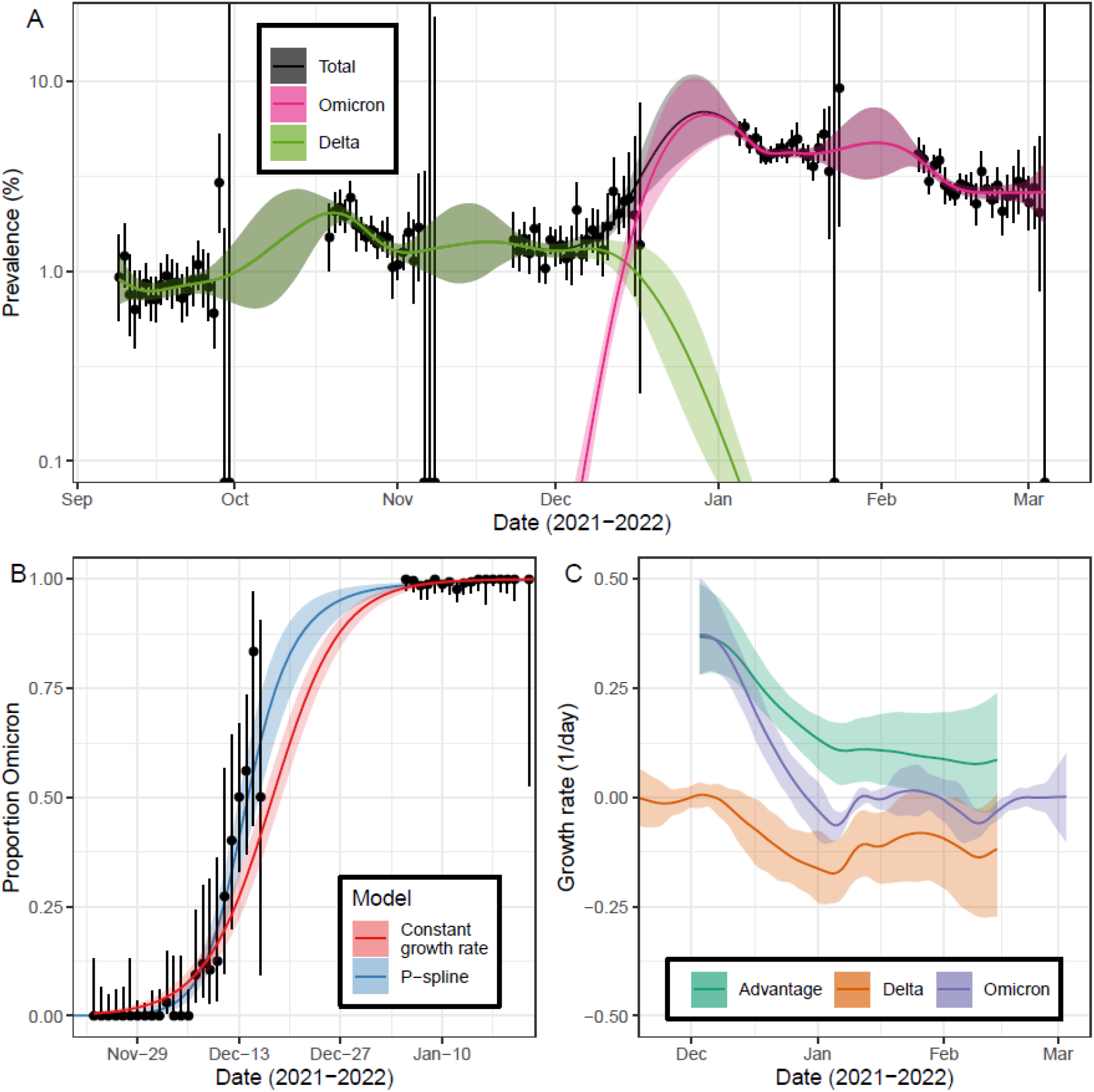
Competition of Omicron and Delta variants. A) Modelled prevalence of SARS-CoV-2 variants Omicron and Delta in England estimated using a mixed-effects Bayesian P-spline model. Estimates of prevalence are shown with a central estimate (solid line) and 95% (shaded region) credible intervals. Daily weighted estimates of prevalence (points) are shown with 95% credible intervals (error bars). B) Modelled proportions of lineages identified as Omicron in England, estimated using Bayesian logistic regression (red) and using a mixed-effects Bayesian P-spline model (blue). Estimates are shown with a central estimate (solid line) and 95% credible intervals (shaded region). Daily estimates of the proportion of lineages Omicron (points) are shown with 95% confidence intervals (error bars). C) Daily growth rate of Omicron (purple), Delta (orange) and their additive difference (green) estimated from the mixed-effects Bayesian P-spline model. Estimates are shown with a central estimate (solid line) and 95% credible intervals (shaded region).

We measured an average daily growth rate in the log-odds of Omicron infection of 0.21 (0.20, 0.23) during rounds 16 to 18 (23 November 2021 – 1 March 2022). However, our results suggest that the growth advantage was not constant over time and ranged from 0.37 (0.28, 0.49) on the 3 December (first day Omicron detected in the REACT-1 study) declining steadily to 0.11 (0.03, 0.17) on 8 January (Fig. 1C). This change in growth advantage over time could be explained by a shorter generation time for Omicron [15] estimated to be approximately 28% shorter than that of Delta [15]. The decline in growth rate may reflect the virus initially achieving higher average rates of transmission among younger and more socially active groups than in the population as a whole [23]. A similar decrease in growth advantage over time was detected when the Alpha variant emerged in England during late 2020 [3].

Models fit to the nine regions of England (Sup Figs. 3-5) and to four broad age-groups (Sup Figs. 6-8) found similar growth advantages in all regions and age-groups. Trends in the proportion of cases that were Omicron showed a high degree of synchrony across regions, but there was some asynchronous behaviour between age-groups. We estimated a 50% proportion Omicron by 10 December (8 December, 14 December) 2021 for 18-34 year olds which was faster than for 5-17 year olds (50% proportion by 25 December (22 December, 29 December) 2021), though this may reflect differences in the prevalence of Delta before Omicron emerged (higher prevalence in 5-17 year olds) (Sup Fig. 6).

As the prevalence of Omicron increased, the prevalence of Delta dropped rapidly to below 0.1% on 3 January 2022 (30 December 2021, 6 January 2022) (Fig.1A) with similar decreases observed in all regions (Sup Fig. 3) and age-groups (Sup Fig. 6). For Delta, we estimated that the time-varying reproduction number (*R*_*t*_) halved in the three weeks from 9 December to 30 December 2021 with values of 1.00 (0.91, 1.10) and 0.50 (0.38, 0.66) respectively (Fig. 2A). The rapid increase in Omicron infections leading to a depletion of the population susceptible to Delta may at least partially explain this reduction in *R*_*t*_. The contribution to that drop of changes in behaviours [24] and public health measures aimed at reducing transmission [25] remains uncertain.

**Figure 2:**
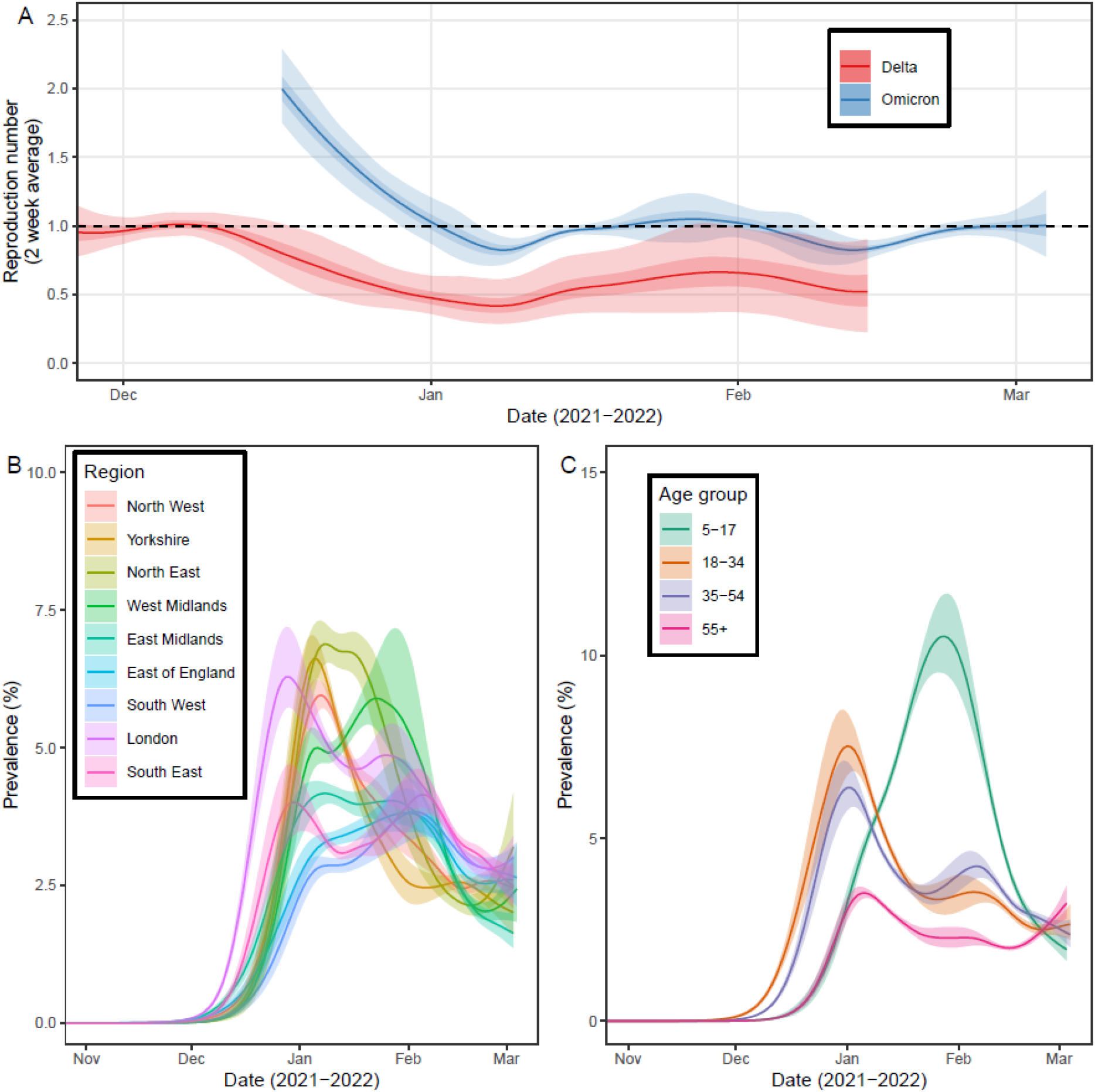
Epidemic dynamics of the Omicron wave. A) Rolling two-week average (prior two weeks) Reproduction number for Omicron and Delta in England as inferred from the mixed-effects Bayesian P-spline model. Estimates are shown with a central estimate (solid line) and 50% (dark shaded region) and 95% (light shaded region) credible intervals. Dashed line shows R=1 the threshold for epidemic growth. B) Modelled prevalence of Omicron in each region of England, estimated using a mixed-effects Bayesian P-spline model. Estimates are shown with a central estimate (solid line) and 50% (shaded region) credible intervals. 95% credible intervals and daily point estimates are included in supplementary figure 1. C) Modelled prevalence of Omicron for four age-groups in England, estimated using a mixed-effects Bayesian P-spline model. Estimates are shown with a central estimate (solid line) and 50% (shaded region) credible intervals. 95% credible intervals and daily point estimates are included in supplementary figure 4.

### Epidemic dynamics of the Omicron wave

The epidemic curve for Omicron swab-positivity had a high initial peak followed by a plateau and possible resurgence (Fig.1A). The national prevalence of Omicron increased rapidly reaching a maximum value of 6.89% (5.34%, 10.61%), on 30 December 2021 (21 December 2021, 31 January 2022) (Sup Fig. 9, Sup Tab. 1). During January prevalence decreased to 4.18% (4.00%, 4.37%) on 12 January where it plateaued with the beginnings of a resurgence detected in late-January 2022 (Fig. 1A). By early-March 2022 prevalence was approximately constant at 2.60% (2.20%, 3.04%) on 1 March (the last official day of round 18 of the study). Trends in the 2-week average instantaneous reproduction number, *R*_*t*_ (Fig.2A), showed that on 17 December 2022 (2 weeks after first Omicron detected) *R*_*t*_ was well above one with a value of 1.99 (1.75, 2.29) despite high levels of vaccine coverage (1 dose 89.8% of those 12 years or older, 2 doses 82.1%, 3 doses 56.9%) [26]. Into early January 2022 *R*_*t*_ rapidly decreased with the central estimate falling below one on 2 January 2022 before rising to above one in late January (21 January to 1 February). Through February 2022 *R*_*t*_ was below one, but at the end of round 18 *R*_*t*_ was no longer securely below one with an *R*_*t*_ value of 1.00 (0.88, 1.12) on 1 March 2022, and a probability of *R*_*t*_ >1 of 0.50.

During December 2021 Omicron’s prevalence rapidly increased in all regions of England (Fig. 2B) though there was heterogeneity in the timing and magnitude of the peak (Sup Fig. 9, Sup Tab. 1). The maximum prevalence reached was highest in the North East at 7.37% (6.42%, 9.79%) and lowest in the East of England at 3.98% (3.38%, 5.80%). Maximum prevalence was reached fastest in London, peaking at 6.45% (5.15%, 10.27%) on 29 December 2021 (25 December 2021, 28 January 2022). In contrast, prevalence reached a maximum of 4.12% (3.21%, 6.36%) in the South West on 3 February (11 January, after 1 March) 2022, approximately 5 weeks later than in London. The rapid rise in prevalence in London could not be explained by a higher regional value of *R*_*t*_ with estimates in December being highly comparable between all regions (Sup Fig. 10) and may therefore be related to an earlier introduction of Omicron in London. Trends over time in regional *R*_*t*_ estimates were comparable across regions and followed a similar pattern as the national estimates. On 1 March central estimates of *R*_*t*_ were above one in 4 regions: North East, East of England, London and West Midlands.

Trends over time in age-group specific *R*_*t*_ also showed a high degree of synchrony (Sup Fig. 11). However, *R*_*t*_ in those aged 5-17 years old was found to be higher in January 2022 relative to other age-groups (although lower in February) leading to a longer initial period of uninterrupted growth whilst *R*_*t*_ for other age-groups dropped below one in early-January 2022. Prevalence in 5-17 year olds (Fig. 2C) peaked on 28 January (21 January, 1 February) 2022, reaching a maximum prevalence of 10.74% (8.52%, 14.74%). This was almost 50% higher than the next highest age-group (18-34 year olds), which had a maximum prevalence of 7.65% (6.08%, 12.35%) approximately 4 weeks earlier on 1 January 2022 (27 December 2021, 5 January 2022) (Sup Fig. 9, Sup Tab. 1). The prevalence was lowest in those aged 55 and over with a maximum prevalence of 3.67% (3.25%, 4.88%) reached on 7 January (1 January, after 1 March) 2022. However, there was indication that prevalence in this group was increasing at the end of the study, with an *R*_*t*_ estimate of 1.14 (0.97, 1.33) on 1 March, a greater estimate than in the other age-groups.

### Omicron sub-lineage competition

The proportion of the Omicron sub-lineage BA.1 decreased over rounds 16, 17 and 18 of REACT-1 (Sup Tab. 2) with the proportion of BA.2 increasing over the same period, and the proportion of BA.1.1 increasing up to 8 February (7 February, 9 February) 2022 (Fig. 3A). On the 30 December 2021 when Omicron’s prevalence reached its maximum the proportion of BA.1 was at 84.6% (82.9%, 86.2%), BA.1.1 at 15.2% (13.6%, 16.9%) and BA.2 at only 0.2% (0.1%, 0.3%). However, by 1 March the proportion of BA.1 was at 9.6% (8.1%, 11.3%), BA.1.1 at 21.6% (18.7%, 24.9%) and BA.2 was now at 68.7% (64.6%, 72.7%).

**Figure 3:**
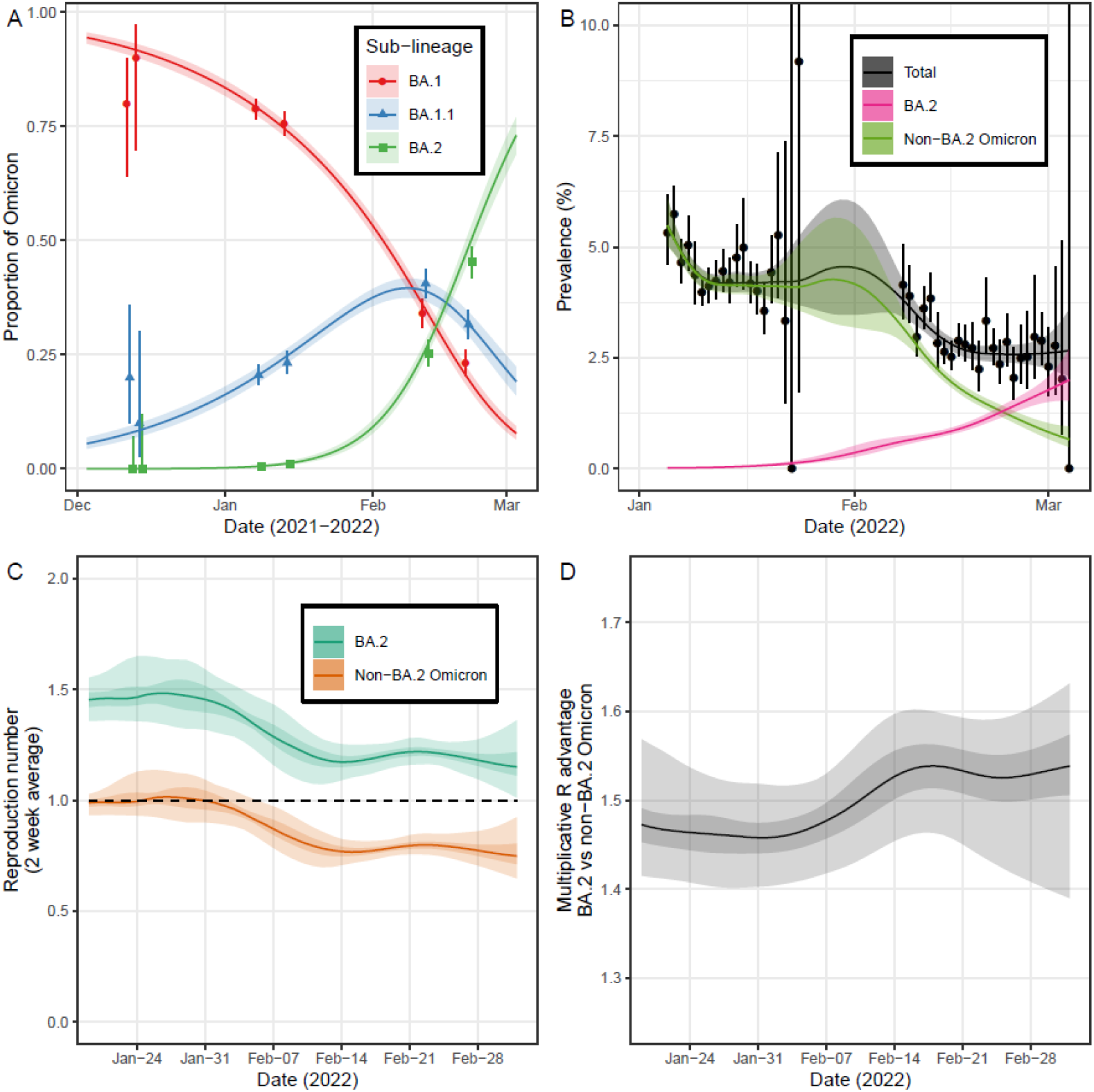
Dynamics of Omicron sub-lineages. A) Modelled proportion of Omicron lineages that are BA.1, BA.1.1 and BA.2, estimated for using a Bayesian multinomial logistic regression model. Estimates of proportion are shown with a central estimate (solid line) and 95% (shaded region) credible intervals. Point estimates of proportion (points) are shown with 95% confidence intervals (error bars) for each half round of REACT-1 with x-axis value set by the median date (jittered for all lineages for visualisation). B) Modelled prevalence of BA.2 and non-BA.2 Omicron in England estimated using a mixed-effects Bayesian P-spline model. Estimates of prevalence are shown with a central estimate (solid line) and 95% (shaded region) credible intervals. Daily weighted estimates of prevalence (points) are shown with 95% credible intervals (error bars). C) Rolling two-week average (prior two weeks) Reproduction number for BA.2 and non-BA.2 Omicron in England as inferred from the mixed-effects Bayesian P-spline model. Estimates are shown with a central estimate (solid line) and 50% (dark shaded region) and 95% (light shaded region) credible intervals. Dashed line shows R=1 the threshold for epidemic growth. D) Multiplicative advantage in the two-week average reproduction number for BA.2 vs non-BA.2 Omicron in England as inferred from the mixed-effects Bayesian P-spline model. Estimates are shown with a central estimate (solid line) and 50% (dark shaded region) and 95% (light shaded region) credible intervals.

The daily log-odds of BA.1.1 infection increased with a daily growth rate of 0.042 (0.037, 0.046) relative to BA.1, while the daily log-odds of BA.2 infection increased with a daily growth rate of 0.133 (0.122, 0.144) relative to BA.1. There was also a difference in the growth between BA.2 and BA.1.1 with a daily growth rate in the log-odds of BA.2 infection of 0.091 (0.081, 0.102) relative to BA.1.1. This shows that the transmissibility of BA.2 and BA.1.1 are both greater than BA.1 but BA.2 has the highest rate of transmission. This increased growth rate of BA.2 in England has also been detected by the UK Health Security Agency (UKHSA) [27] and multiple other countries have observed an increasing proportion of BA.2. Increasing proportions of BA.1.1 relative to BA.1 have been observed in Denmark [28], and in the USA BA.1.1 was the major Omicron variant during Omicron’s initial emergence [29] suggesting BA.1 was outcompeted before it could establish there.

In England we estimated different trends in the national prevalence of BA.2 vs non-BA.2 Omicron infections over the period of rounds 17 to 18 (Fig. 3B, Sup Fig. 12) under the assumption that all infections were Omicron (<1% were non-Omicron infections in both rounds). During February 2022, the prevalence of BA.2 steadily increased, whereas the prevalence of non-BA.2 Omicron decreased. An increasing prevalence of samples that are positive on the S-gene (an approximate proxy for BA.2) was reported in England over the same period [30]. Consistently, the estimated *R*_*t*_ for BA.2 was greater than that of non-BA.2 Omicron (Fig. 3C) with an estimate of 1.17 (1.08, 1.28) for BA.2 and 0.77 (0.69, 0.87) for non-BA.2 Omicron on 1 March. The difference in *R*_*t*_ over time corresponded to a multiplicative advantage for BA.2 over non-BA.2 Omicron of approximately 1.5 (Fig. 3D), with daily estimates ranging between 1.46 (1.40, 1.52) on 31 January 2022 and 1.54 (1.46, 1.60) on 18 February 2022. This difference in dynamics between Omicron sub-lineages can explain the observed national trends in Omicron prevalence, with *R*_*t*_ increasing towards late February 2022 due to the rising proportion of BA.2. We find that a greater proportion of BA.2 infected individuals exhibit the most predictive COVID-19 symptoms (loss or change of sense of smell or taste, fever, new persistent cough) compared to those infected with BA.1 (Sup Tabs. 3-4).

Analogous models fit to data subset by region (Sup Figs. 13-15) and age-group (Sup Figs 16-18) showed similar growth rate advantages for BA.2 for all age-groups but a small degree of heterogeneity between regions; the advantage was greater in East of England relative to the South West (all other regions were comparable). Higher proportions of BA.2 were reported in London and the South East [27]; as we do not estimate a higher growth advantage in these two regions the greater proportion of BA.2 suggests earlier introductions there, in agreement with the higher rates of international travel from these regions [31]. Phylogeographic analysis further supported this with London being highly represented in estimates for the regions of many ancestral nodes within Omicron’s phylogenetic tree (Sup Fig. 19). However, during periods in which the Omicron sub-lineages were well-sampled there was little geographic structure present in Omicron’s phylogeny. This is potentially due to the rapid time-frame by which each Omicron sub-lineage in turn was disseminated across the country following their introduction in London. We estimated higher symmetrical region-to-region migration rates from London to other regions over all rounds and for each Omicron sub-lineage (Sup Tab. 5); they were most consistently high for London to South East and London to North West.

The estimated proportion of BA.2 over time was similar in all age-groups (Sup Fig. 17) and so could not explain the higher estimates of *R*_*t*_ in those aged 55 and over in late-February/early-March 2022. Central estimates for the *R*_*t*_ of BA.2 were greater than one on 1 March in all regions (Sup Fig. 20) and age-groups (Sup Fig. 21) and so as the proportion of BA.2 further increases a resurgence in prevalence would be expected. A resurgence has been observed across all regions and age-groups in the numbers of cases and hospitalisations recorded in the routine data during the first three weeks of March 2022 [26]. The emergence of BA.2 has acted to prolong the Omicron wave of the epidemic in England.

### Limitations

Our study has limitations. The sampling is performed over discrete rounds with periods of no data, for which trends have to be inferred. One such period was late December 2021, a key period of growth in the proportion of SARS-CoV-2 infections caused by the Omicron variant, leading to wide credible intervals for the dynamics of the pandemic over this period. Sequencing is unlikely to be successful on samples with a low viral load and so was only performed on samples with an N-gene cycle threshold (Ct) value (a proxy for viral load) less than 34. However, it is unknown if there are intrinsic differences in the Ct values by lineage which could bias estimated proportions. Further, differences in Ct values due to differences in growth rate [32] could also lead to more transmissible variants being detected more favourably.

Finally, our study estimates the daily prevalence, the proportion of the population testing positive, and not the incidence. This can lead to estimates of *R*_*t*_ being overly smoothed due to swab-positivity remaining over several days [33,34], and trends of *R*_*t*_ over time may be lagged by a period depending on the duration for which individuals test positive. It is also worth noting that our estimates of *R*_*t*_ are based on specific estimates of the generation time distribution; studies using different generation time distributions will return different estimates of *R*_*t*_ [35]. Additionally our region- and age-group-specific *R*_*t*_ estimates assume that all infections in a particular subgroup result from contact/mixing among members of that subgroup and so, though highly informative, must be interpreted cautiously.

## Conclusion

As the cumulative incidence and vaccination coverage continue to increase, the SARS-CoV-2 virus will find itself competing against a diverse and complex immunity landscape within the human population. Accordingly, the evolutionary dynamics of the virus will be dominated by immune evasion. This has already been observed with the emergence of the Omicron variant and its sub-lineages, the consequence of which was an initial wave of infection peaking at a prevalence of 6.89% in England, the highest recorded at any time hitherto in the REACT-1 study. These infection rates occurred against a background of high levels of vaccine coverage and past infections, further fuelled by the emergence of the more transmissible Omicron sub-lineage BA.2. Given the regular emergence of VOCs during the first two years of the COVID-19 pandemic there is little reason to believe this trend will not continue. Indeed, other respiratory infections such as Influenza observe annual epidemics due to the emergence of new strains better able to navigate the immune landscape [36,37]. If we see a similar trend for SARS-CoV-2 then intermittent waves of infection of a similar magnitude to Omicron are within the bounds of possibility. Continued surveillance, booster vaccinations and, potentially, updates of the vaccines will be crucial in minimising the harmful effects of this new public health paradigm. Greater vaccine equity worldwide can help reduce the rate at which these harmful variants emerge [38].

## Methods

### REACT-1 study protocol

The methodology of REACT-1 has been described in detail elsewhere [39]. In short, each round a random subset of the population in England is selected at the lower tier local authority (LTLA) level (N=315) and invited to participate in the study. Those who agree to participate provide a self-administered (parent/guardian administered for those aged 5-12 years old) throat and nose swab which undergoes rt-PCR testing for the SARS-CoV-2 virus. Individuals are classified as positive if their test has an N-gene Ct value less than 37 or if both the N- and E-gene are detected. Rim weighting [40] is used to weight individual test results by age, sex, deciles of the Index of Multiple Deprivation, LTLA counts, and ethnic group. Analysis was performed using rounds 14 to 18 of the study running from 9 September 2021 to 1 March 2022. During rounds 15 to 18 of the study all swab tests were sent to the lab via the post, whereas in round 14 of the study approximately 50% of tests were collected via courier. No difference in samples were observed for the two different collection methods [41].

### Sequencing

All swab tests with an N-gene Ct value less than 34 and sufficient volume underwent genomic sequencing. Extracted RNA was amplified using the ARTIC protocol [42] with sequence libraries provided by CoronaHiT [43]. Sequencing was performed on the Illumina NextSeq 500 platform. Analysis of the raw sequencing was done using the bioinformatics pipeline[44] before being uploaded to CLIMB [45]. Lineage designation was then performed using PangoLEARN [46] (database version 2022-02-28), a machine-learning based algorithm for lineage designation which uses the PANGO nomenclature [5]. Sequences could not be obtained for some samples of low overall quality. Further, samples for which at least 50% of bases were not covered were excluded from the analysis.

### Mixed-effects Bayesian P-spline model

A mixed-effects Bayesian P-spline model was used to estimate the prevalence of both Omicron and Delta SARS-CoV-2 infections over time. The basic Bayesian P-spline model has been described in detail [47]. In short, the entire time series is split into equally sized knots (approximately 5 days apart) with 3 further knots defined at both the beginning and end of the time series (to prevent edge effects). A system of 4th order basis-splines (b-splines) is defined over all knots. The P-spline for a single lineage’s prevalence is then defined as a linear combination of these basis splines:

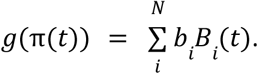

Here *g*() is the logit link function, π(*t*) is the prevalence on day *t, b*_*i*_ are the b-spline coefficients, and *B*_*i*_ (*t*) is the value of the *i*^*th*^ b-spline on day *t*. A second-order random-walk prior distribution is defined for the b-spline coefficients, *b*_*i*_ = 2*b*_*i*−1_− *b*_*i*−2_ + *u*_*i*_, where *u*_*i*_ ∼*N*(0, ρ). The first two coefficients, *b*_1_ and *b*_2_, are given uninformative constant prior distributions. The parameter ρ controls the smoothness of the curve and penalises changes in the first derivative (approximately the growth rate) reflecting the prior knowledge we have of an epidemic system. P-splines are defined for the prevalence of both Omicron and Delta with ρ being a shared parameter. A further prior distribution taking the form *u*_*i,Omicron*_ − *u*_*i,Delta*_ ∼*N*(0, η) is defined on the changes in the first derivative for both lineages. The parameters ρ and η were both given uninformative inverse gamma prior distributions η, ρ ∼ *IG*(0. 001, 0. 001). This assumes that changes in the growth rate happen simultaneously for both Omicron and Delta, which effectively assumes a constant growth rate advantage, unless there is significant evidence to the contrary.

The sum total of Omicron and Delta’s modelled prevalence is fitted to the daily weighted number of tests and positive tests assuming a binomial likelihood. Simultaneously the proportion of the total prevalence attributed to Omicron is fitted to the daily number of Omicron lineages vs total number of samples with a lineage determined, again assuming a binomial likelihood. The model is fit to rounds 14 to 18 of the REACT-1 data using a No-U-Turns sampler [48] implemented in STAN [49]. Models are also fit to the data subset by region of England, and subset by age quartile. An analogous model is fit to rounds 17 and 18 instead comparing BA.2 and non-BA.2 Omicron lineages under the assumption that total prevalence was caused only by Omicron lineages (>99% Omicron in both rounds).

Estimates of the instantaneous growth rate of Omicron, Delta and their difference were estimated over time from the modelled prevalence time-series. The rolling two-week average reproduction number for both Omicron and Delta was also estimated from the modelled prevalence using methodology that has previously been described [47]. The model assumed a gamma-distributed generation time with rate parameter = 0.27, and shape parameter = 0.89, for Omicron or Omicron sub-lineages (BA.2, non-BA.2 Omicron lineages) and rate parameter = 0.48 and shape parameter = 2.20 for Delta [15].

### Constant growth rate models

The daily growth rate in the log-odds of Omicron infection relative to Delta infection, assuming a constant growth rate for the whole period, was estimated using a Bayesian logistic regression model fit to a binary outcome variable (Omicron or Delta) over time. The daily growth rate in the log-odds of BA.2 and BA.1.1 relative to BA.1 over rounds 17 and 18, assuming constant growth rates, was estimated using a Bayesian multinomial logistic regression model fit to the categorical outcome variable (BA.1, BA.1.1 or BA.2) with BA.1 set as the reference category. The difference in these two growth rates was used to estimate the daily growth rate in the log-odds of BA.2 relative to BA.1.1.

### Phylogeographic analysis

Phylogeographic analysis was performed on lineages that were designated as Omicron (BA.1, BA.1.1, BA.2). A maximum likelihood phylogenetic tree assuming a HKY model was fitted to the sequences using IQTree [50]. A relaxed molecular clock model, assuming a mean evolutionary rate of 0.0008 substitutions/site/year, was fit to the tree using TreeTime [51] to give a time-resolved phylogenetic tree. A further mugenic model again implemented in TreeTime [51] was fit to the time-resolved phylogeny treating the region of England (N=9) where each sample was obtained as a discrete state. From this model we estimated the mean pairwise migration rate of Omicron between all regions of England. We excluded sequences without complete date information since they do not contribute to the estimation of divergence times. Sequences with an excess of gaps cannot be placed in the phylogeny correctly, and so we excluded sequences with less than 75% of bases covered. Note that this is a more stringent threshold than was used earlier for the task of lineage classification which can be performed for sequences with fewer bases covered. We further excluded one sequence which deviated too much from a preliminary strict clock (more than five times the interquartile range from the clock regression).

Omicron sequences with at least 75% of bases covered were compared to all sequences deposited in GISAID after the 2nd Dec 2021, with the 500 closest neighbours extracted using uvaiann [52]. The similarity measure is based on the number of single nucleotide polymorphism (SNP) matches, number of partial matches, and number of valid comparisons so that we prefer more resolved sequences. Matches to GISAID samples from the REACT-1 study were excluded afterwards. We then compared the number of samples that match (no different SNPs) a REACT-1 sequence by the country where they were collected. We additionally investigated the number of samples that were at specific SNP distance (1,2,3 and 4 SNP mismatches) from REACT-1 sequences again by the country where they were collected. The reported number of SNP mismatches considers partially ambiguous DNA codes: DNA bases which cannot be unambiguously inferred by the assembler may be reported as e.g. ‘M’ to indicate that the base may be an adenine (A) or a cytosine (C) [53]. Such a state is compatible and thus considered a match to another sequence which has for instance an ‘A’ in the same genomic location.

In order to estimate potential Omicron importations into and exportations from England based on the REACT-1 samples, we used the set of closest neighbours described above, restricting to global sequences sampled within one week of REACT-1 samples with at most one SNP distance. We furthermore removed global GISAID matches from the UK (i.e. sequences deposited in GISAID from the UK), and for each REACT-1 sequence we kept only the global match with the highest number of valid pair comparisons (locations where neither sequence is a gap or have low coverage). After removing duplicate hits, since the same global reference can be the best match for more than one REACT-1 sequence, we inferred a potential importation if the sampling date of the global sequence is earlier than its matching REACT-1 sample, and as a potential exportation if the REACT-1 sample is earlier (but still within one week). Global sequences which matched both an earlier and a later REACT-1 sample were removed (since they could be inferred as a source or destination of the migration). In total we have 335 imported samples and 310 exported ones. The date of the importation/exportation was taken as the date of the second sample for each pair. The actual import/export dates may be earlier due to an importation lag [54].

### Statistical analyses

The Wilson method [55] which is prefered for low numbers of positives [56] was used to calculate the 95% confidence intervals for all lineage proportions.

The proportion of individuals reporting any symptoms, and the proportion reporting the most predictive COVID-19 symptoms (loss or change of sense of smell or taste, fever, new persistent cough) [57] in the last month was estimated in round 16 for those infected with Omicron and Delta, and in rounds 17-18 for those infected with BA.1, BA.1.1 and BA.2. The combination of lineages and rounds chosen was done to avoid introducing biases due to changing rates of symptoms over time, and to ensure a large enough sample of each lineage for calculations to be meaningful. P-values for differences in the proportion reporting symptoms between lineages was estimated using logistic regression models with symptom status (any symptom vs no symptoms and separately most predictive COVID-19 symptoms vs not reporting the most predictive COVID-19 symptoms) as the outcome variable. The sensitivity of any result that was significant (P-value <0.05) was assessed using multivariable logistic regression models including round of the study and N-gene Ct value as additional covariates.

## Supporting information

Acknowlegdements of gisaid contributing authors

## Data Availability

Access to REACT-1 individual-level data is restricted to protect participants' anonymity.
Summary statistics, descriptive tables, and code from the current REACT-1 study are available at https://github.com/mrc-ide/reactidd (doi 10.5281/zenodo.6242826). REACT-1 study materials are available for each round at
https://www.imperial.ac.uk/medicine/research-and-impact/groups/react-study/react-1-study-materials/
Sequence read data are available without restriction from the European Nucleotide Archive at https://www.ebi.ac.uk/ena/browser/view/PRJEB37886, and consensus genome sequences are available from the Global initiative on sharing all influenza data (GISAID).

## Ethics

We obtained research ethics approval from the South Central-Berkshire B Research Ethics Committee (IRAS ID: 283787).

## Contributors

PE, CAD, and MC-H are corresponding authors. OE, SR, PE, MC-H and CAD conceived the study and the analytical plan. OE, LOM, AP performed the statistical analyses. OE, LOM, AP, HWang, BB, DT, DH, and JJ curated the data. CA, WB, GT, GC, HWard, AD provided study oversight and results interpretation. All authors revised the manuscript for important intellectual content and approved the submission of the manuscript. OE, LOM, AP, PE, MC-H, CAD had full access to the data and take responsibility for the integrity of the data and the accuracy of the data analysis and for the decision to submit for publication.

## Declaration of interests

Prof. Elliott is the director of the MRC Centre of Environment and Health (MR/L01341X/1 and MC/S019669/1) and the NIHR Health Protection Research Unit in Chemical and Radiation Threats and Hazards, and has no conflict of interest to disclose. Prof M Chadeau-Hyam holds shares in the O-SMOSE company and has no conflict of interest to disclose. Consulting activities conducted by the company are independent of the present work. All other authors have no conflict of interest to disclose.

## Funding

The study was funded by the Department of Health and Social Care in England. The funders had no role in the design and conduct of the study; collection, management, analysis, and interpretation of the data; and preparation, review, or approval of this manuscript.

## Acknowledgements

AJP acknowledges the support of the Biotechnology and Biological Sciences Research Council (BB/R012504/1). HWard acknowledges support from a National Institute for Health Research (NIHR) Senior Investigator Award, the Wellcome Trust (205456/Z/16/Z), and the NIHR Applied Research Collaboration (ARC) North West London. GC is supported by an NIHR Professorship. PE is Director of the Medical Research Council (MRC) Centre for Environment and Health (MR/L01341X/1, MR/S019669/1). PE acknowledges support from Health Data Research UK (HDR UK); the NIHR Imperial Biomedical Research Centre; NIHR Health Protection Research Units in Chemical and Radiation Threats and Hazards, and Environmental Exposures and Health; the British Heart Foundation Centre for Research Excellence at Imperial College London (RE/18/4/34215); and the UK Dementia Research Institute at Imperial College London (MC_PC_17114). SR and CAD acknowledge support from the MRC Centre for Global Infectious Disease Analysis. CAD acknowledges support from the NIHR Health Protection Research Unit in Emerging and Zoonotic Infections and the NIHR-funded Vaccine Efficacy Evaluation for Priority Emerging Diseases (PR-OD-1017-20007). MC-H and BB acknowledge support from Cancer Research UK, Population Research Committee Project grant ‘Mechanomics’ (grant No 22184 to MC-H). MC-H acknowledges support from the H2020-EXPANSE (Horizon 2020 grant No 874627) and H2020-LongITools (Horizon 2020 grant No 874739).

We thank The Huo Family Foundation for their support of our work on COVID-19. We thank key collaborators on this work – Ipsos MORI: Kelly Beaver, Sam Clemens, Gary Welch, Nicholas Gilby, Kelly Ward, Galini Pantelidou and Kevin Pickering; Institute of Global Health Innovation at Imperial College London: Gianluca Fontana, Justine Alford; School of Public Health, Imperial College London: Eric Johnson, Rob Elliott, Graham Blakoe; Quadram Institute, Norwich, UK: Nabil-Fareed Alikhan; North West London Pathology and Public Health England (now UKHSA) for help in calibration of the laboratory analyses; Patient Experience Research Centre at Imperial College London and the REACT Public Advisory Panel; NHS Digital for access to the NHS register; the Department of Health and Social Care for logistic support.

We thank GISAID for providing global SARS-CoV-2 sequences and gratefully acknowledge all authors (see supplementary materials ‘gisaid_acknowledgements.xlsx’) from the originating laboratories responsible for obtaining the specimens and the submitting laboratories where genetic sequence data were generated and shared via the GISAID Initiative.

## Data availability

Access to REACT-1 individual-level data is restricted to protect participants’ anonymity.

Summary statistics, descriptive tables, and code from the current REACT-1 study are available at https://github.com/mrc-ide/reactidd (doi 10.5281/zenodo.6242826). REACT-1 study materials are available for each round at https://www.imperial.ac.uk/medicine/research-and-impact/groups/react-study/react-1-study-materials/

Sequence read data are available without restriction from the European Nucleotide Archive at https://www.ebi.ac.uk/ena/browser/view/PRJEB37886, and consensus genome sequences are available from the Global initiative on sharing all influenza data (GISAID).

## Supplementary Materials

### Supplementary tables

All supplementary tables are available in the supporting document ‘SupplementaryTables.xlxs’

**Supplementary Table 1:** Estimated maximum prevalence and date of maximum prevalence for all data, and sub-groups by region and age-groups

**Supplementary Table 2:** Number and proportion of each variant for rounds 16, 17 and 18

**Supplementary Table 3:** Number and proportion reporting symptoms by lineage for round 16 (Omicron and Delta) and rounds 17-18 (BA.1, BA.1.1 and BA.2)

**Supplementary Table 4:** Multivariable logistic models investigating the significant differences in symptoms between BA.1 vs BA.2 and BA.1.1 when including round and N-gene Ct value as additional covariates in the model

**Supplementary Table 5:** Average inter-region migration rates, inferred from a mugenic model run on the time-resolved phylogenetic tree for Omicron presented for all samples by round and by Omicron sub-lineage.

### Supplementary figures

**Supplementary Figure 1:**
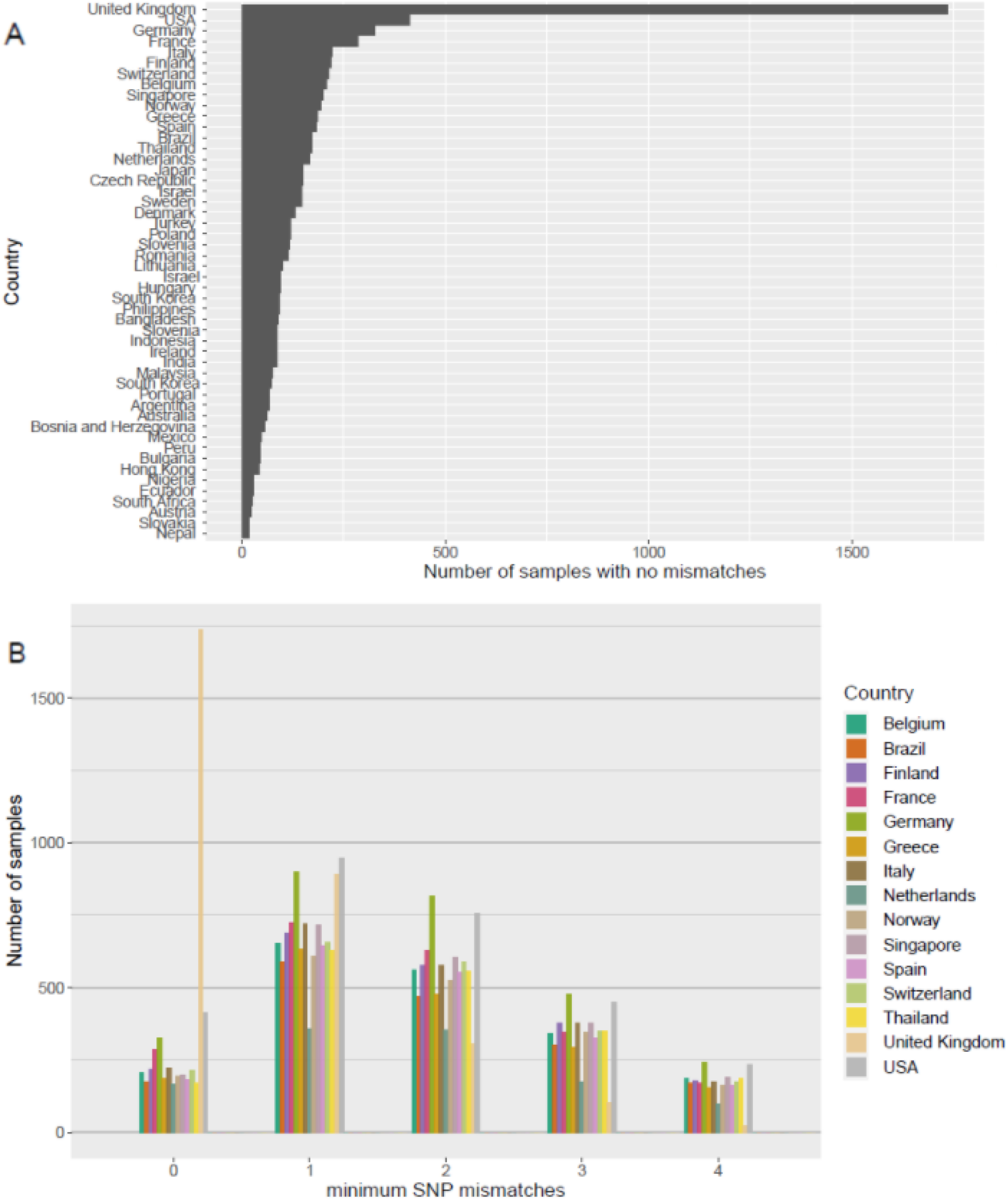
Similarity of REACT-1 sequences to international sequences. (A) The number of REACT-1 sequences that match (no different SNPs) a sequence obtained from GISAID by the country the sequence was sampled in. The top 50 most represented countries have been shown. (B) The number of REACT-1 sequences at a given SNP distant from sequences obtained from GISAID by the country the sequence was sampled in. The top 15 most represented countries have been shown.

**Supplementary Figure 2:**
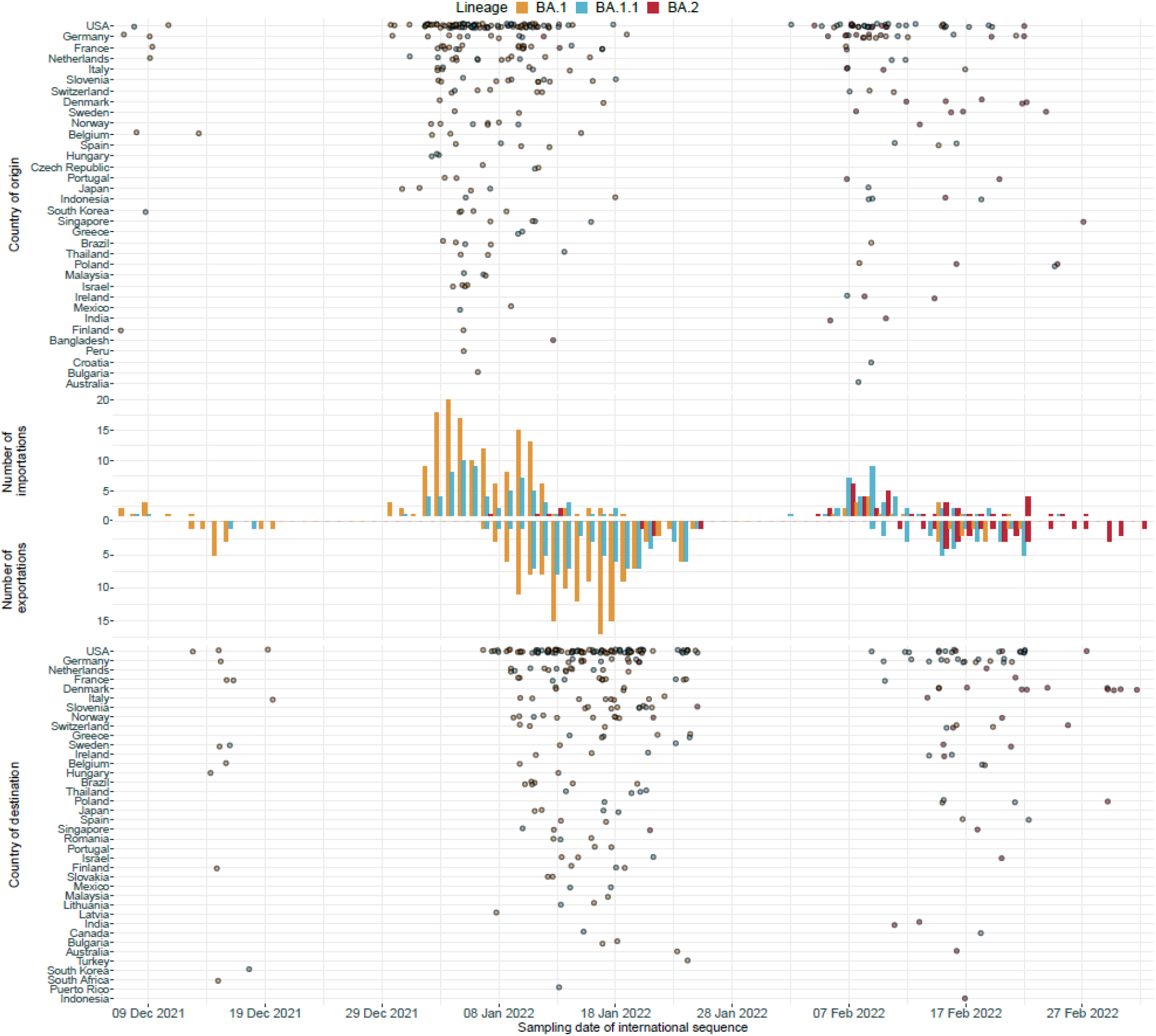
Importation and exportations of Omicron. The daily number of potential importations/exportations (bars) of Omicron to/from England, as inferred with REACT-1 samples and a representative selection of global samples, for BA.1 (orange), BA.1.1 (blue) and BA.2 (red). For each importation/exportation the date of the importation/exportation and the country of the global sample (and thus the origin/destination of the Omicron strain) has been shown (dots).

**Supplementary Figure 3:**
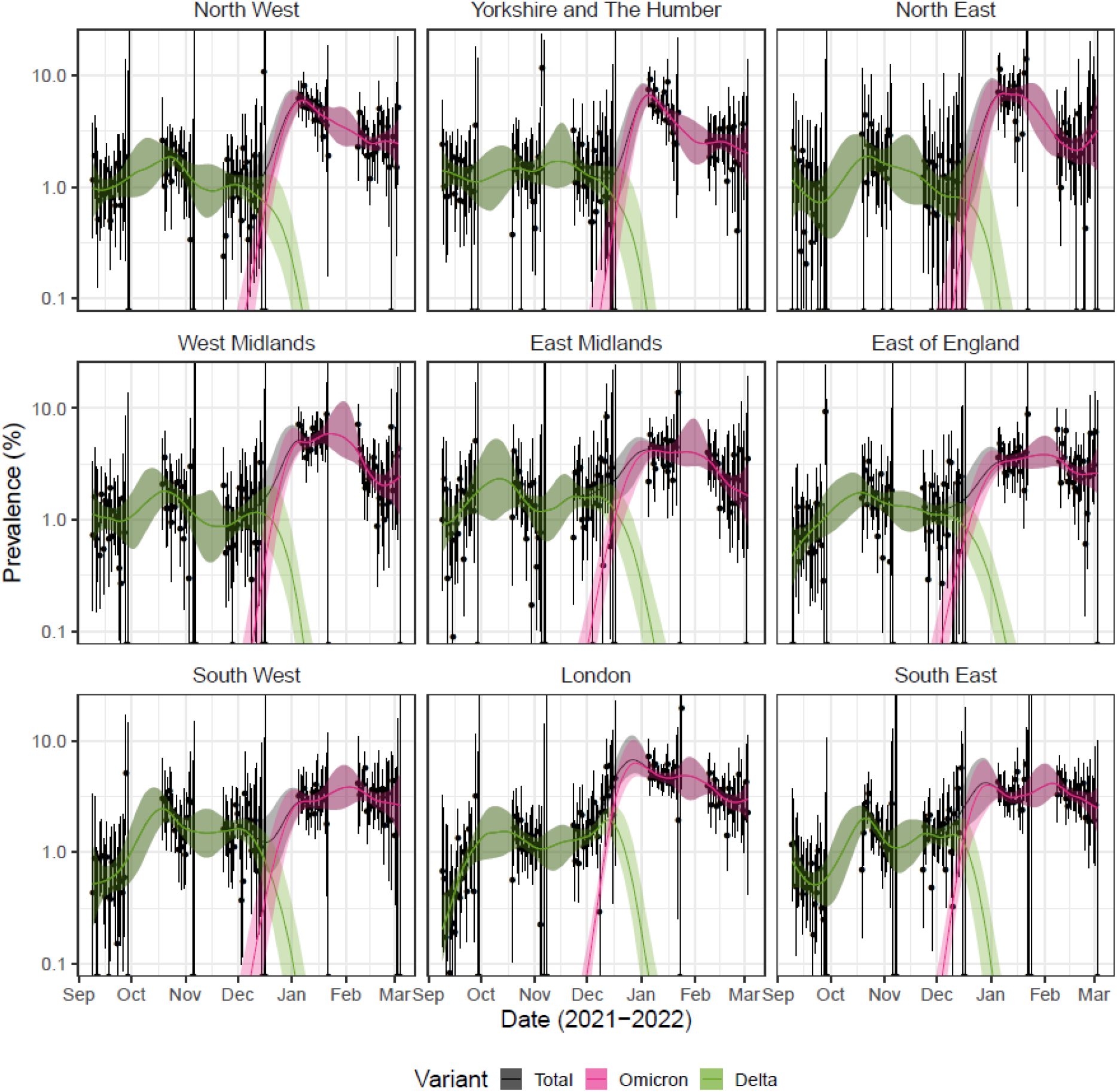
Omicron vs Delta prevalence by region. Modelled prevalence of SARS-CoV-2 variants Omicron (pink) and Delta (green), and total prevalence (grey) in each region of England estimated using mixed-effects Bayesian P-spline models. Estimates of prevalence are shown with a central estimate (solid line) and 95% (shaded region) credible intervals. Daily weighted estimates of prevalence (points) are shown with 95% credible intervals (error bars).

**Supplementary Figure 4:**
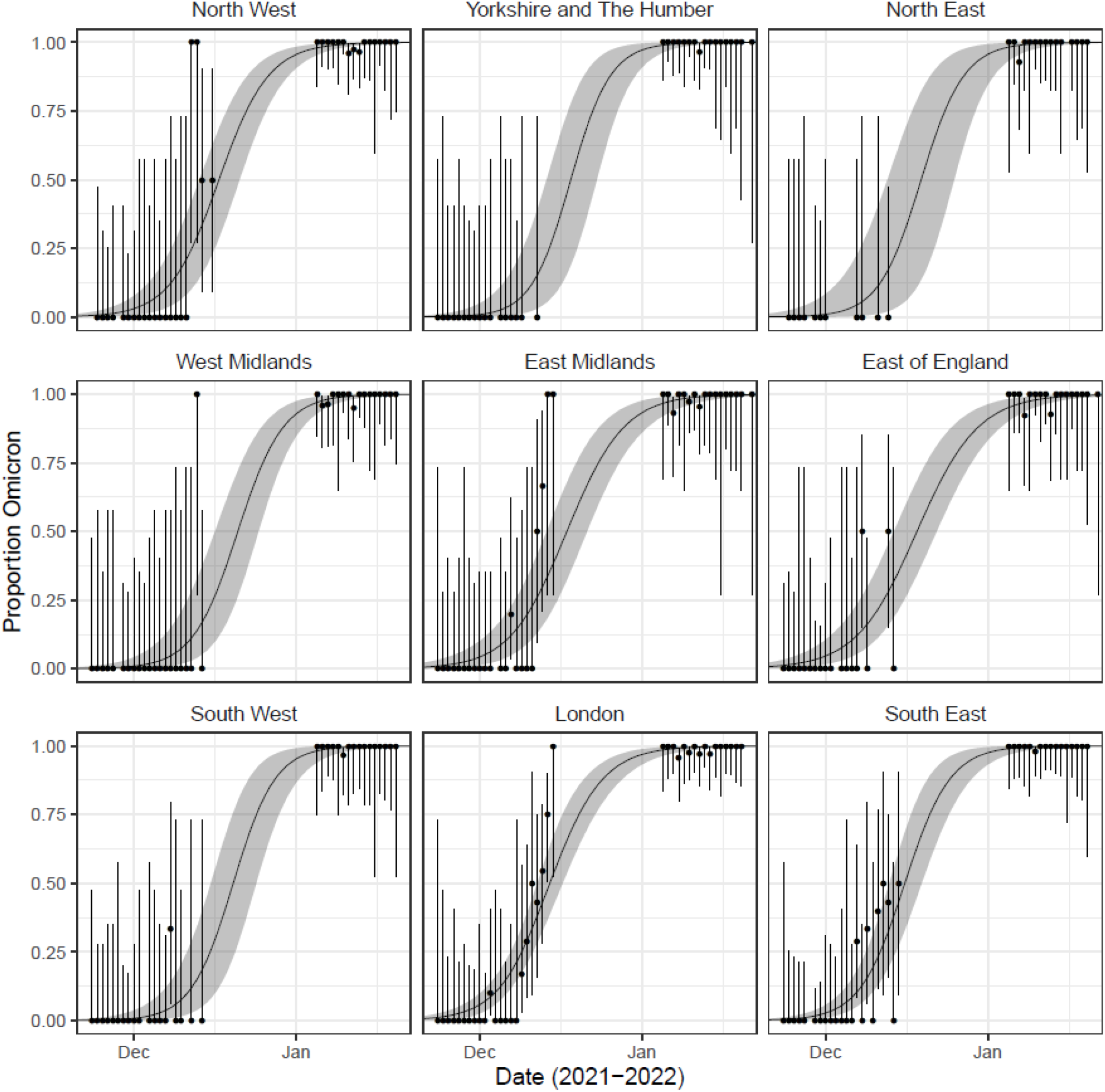
Omicron vs Delta proportion by region. Modelled proportions of lineages identified as Omicron in each region of England estimated using mixed-effects Bayesian P-spline models. Estimates are shown with a central estimate (solid line) and 95% credible intervals (shaded region). Daily estimates of the proportion of lineages Omicron (points) are shown with 95% confidence intervals (error bars).

**Supplementary Figure 5:**
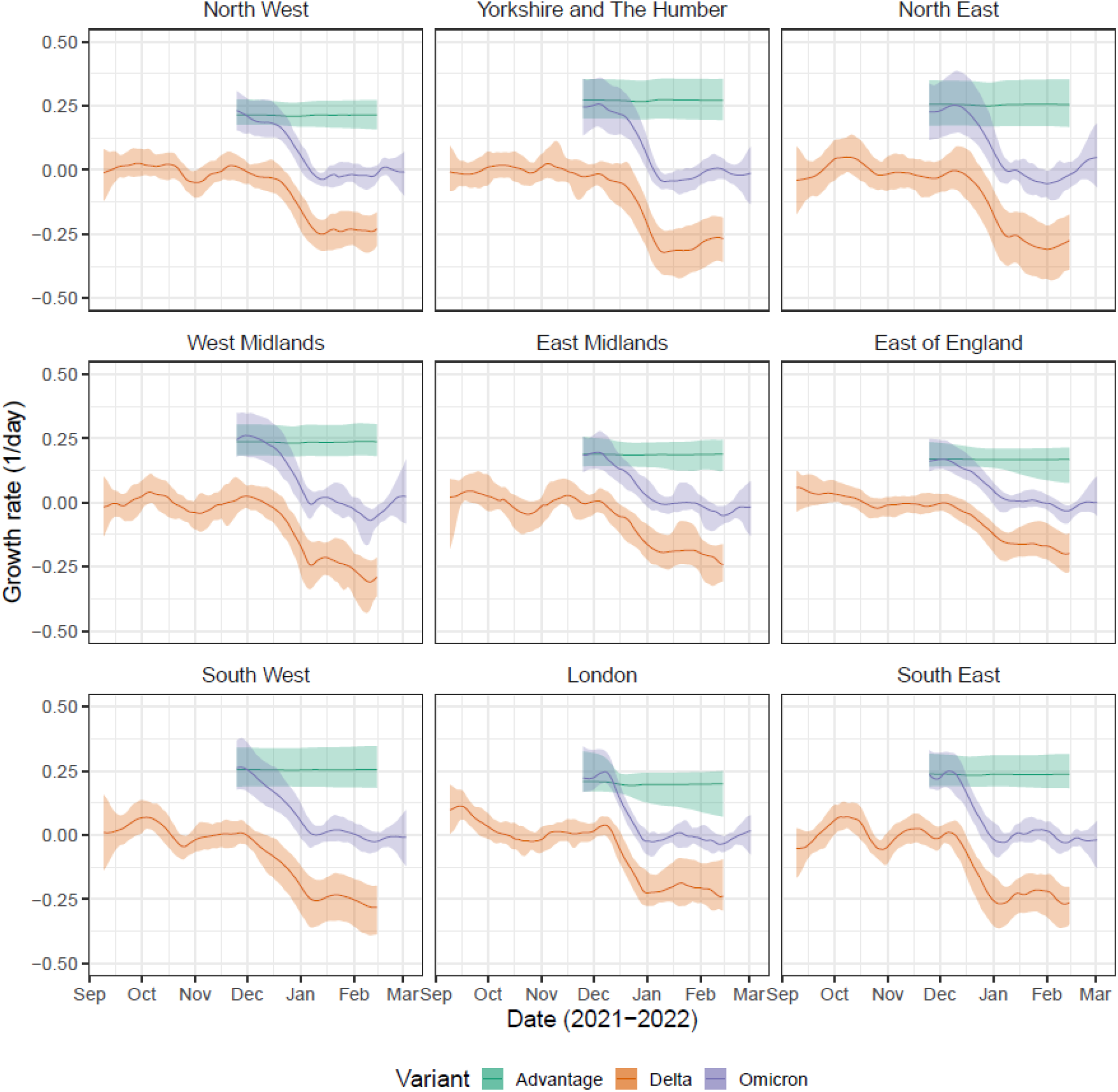
Omicron vs Delta growth rate by region. Daily growth rate of Omicron (purple), Delta (orange) and their additive difference (green) estimated from mixed-effects Bayesian P-spline models fitted to each region of England. Estimates are shown with a central estimate (solid line) and 95% credible intervals (shaded region). Estimates for each lineage are only displayed for the period over which the lineage was detected in REACT-1 samples.

**Supplementary Figure 6:**
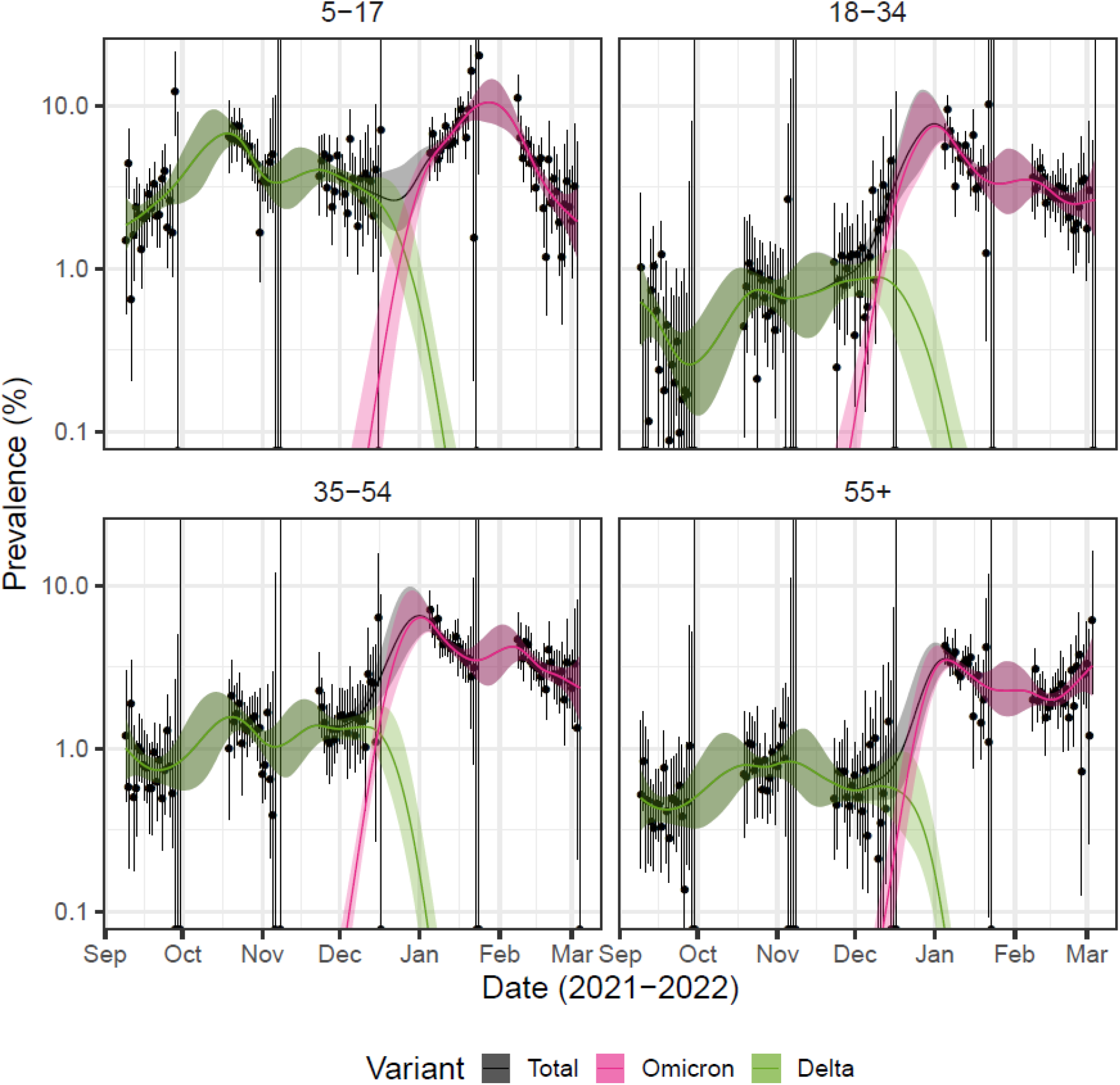
Omicron vs Delta prevalence by age-group. Modelled prevalence of SARS-CoV-2 variants Omicron (pink) and Delta (green), and total prevalence (grey) for each age-group in England estimated using mixed-effects Bayesian P-spline models. Estimates of prevalence are shown with a central estimate (solid line) and 95% (shaded region) credible intervals. Daily weighted estimates of prevalence (points) are shown with 95% credible intervals (error bars).

**Supplementary Figure 7:**
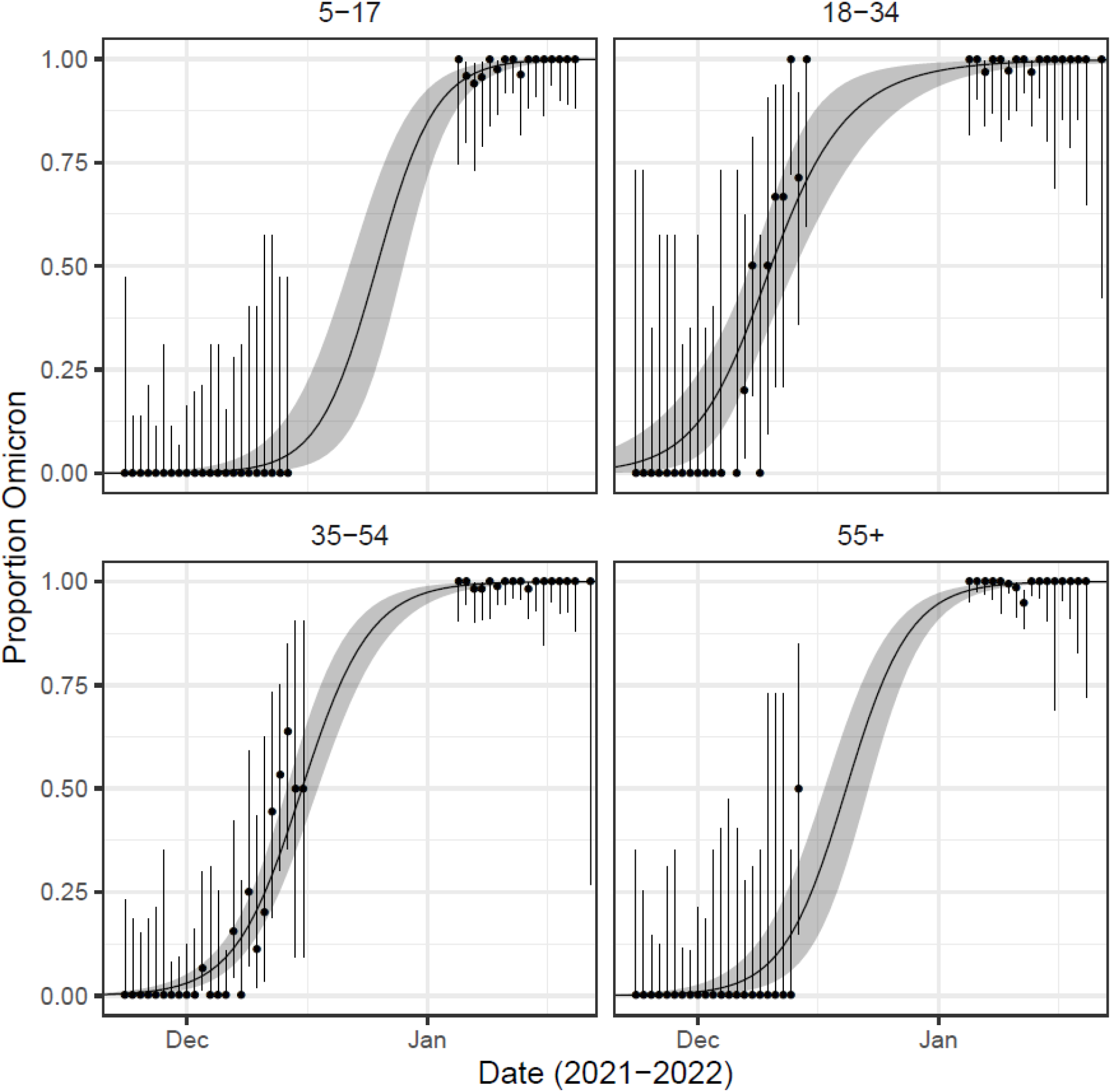
Omicron vs Delta proportion by age-group. Modelled proportions of lineages identified as Omicron for each age-group in England estimated using mixed-effects Bayesian P-spline models. Estimates are shown with a central estimate (solid line) and 95% credible intervals (shaded region). Daily estimates of the proportion of lineages Omicron (points) are shown with 95% credible intervals (error bars).

**Supplementary Figure 8:**
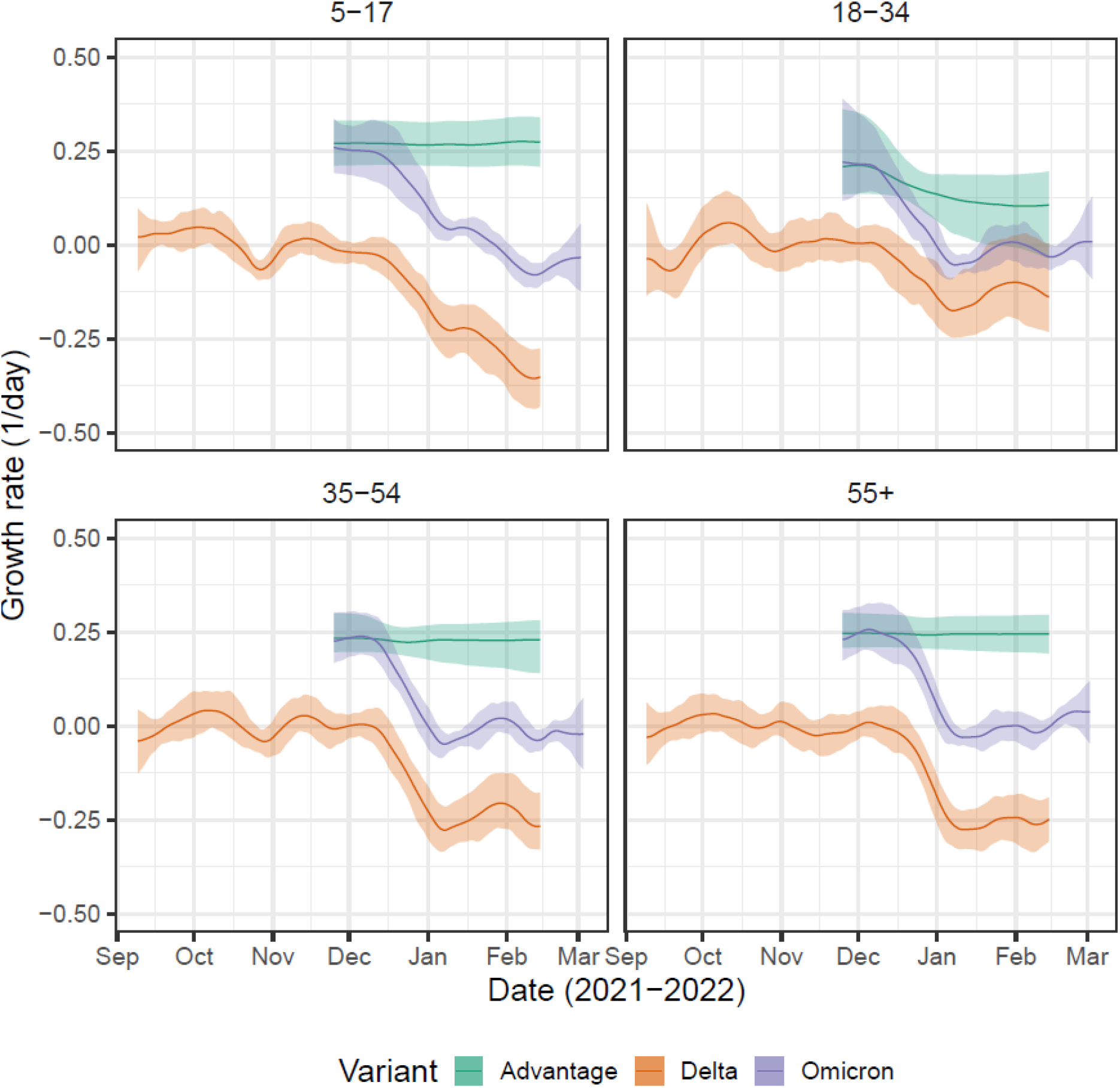
Omicron vs Delta growth rate by age-group. Daily growth rate of Omicron (purple), Delta (orange) and their additive difference (green) estimated from mixed-effects Bayesian P-spline models fitted to each age-group in England. Estimates are shown with a central estimate (solid line) and 95% credible intervals (shaded region). Estimates for each lineage are only displayed for the period over which the lineage was detected in REACT-1 samples.

**Supplementary Figure 9:**
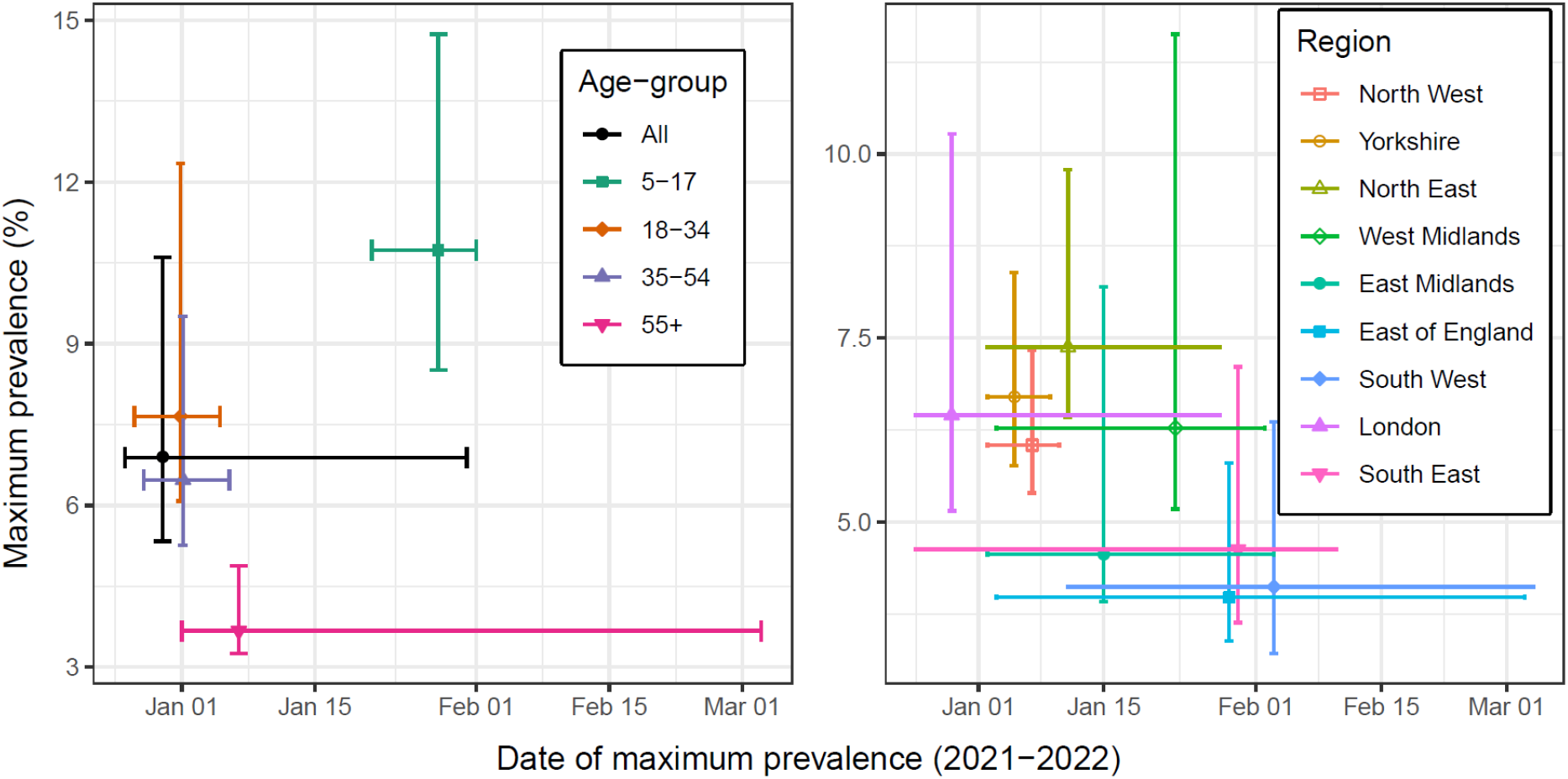
Maximum prevalence and date of maximum prevalence. The maximum prevalence of Omicron reached and the date at which it was reached inferred from the mixed-effects Bayesian P-spline model fitted to all data, and for the models fit to subsets of data by region and age-group. Estimates are shown for the median value (points) with 95% credible intervals (error-bars).

**Supplementary Figure 10:**
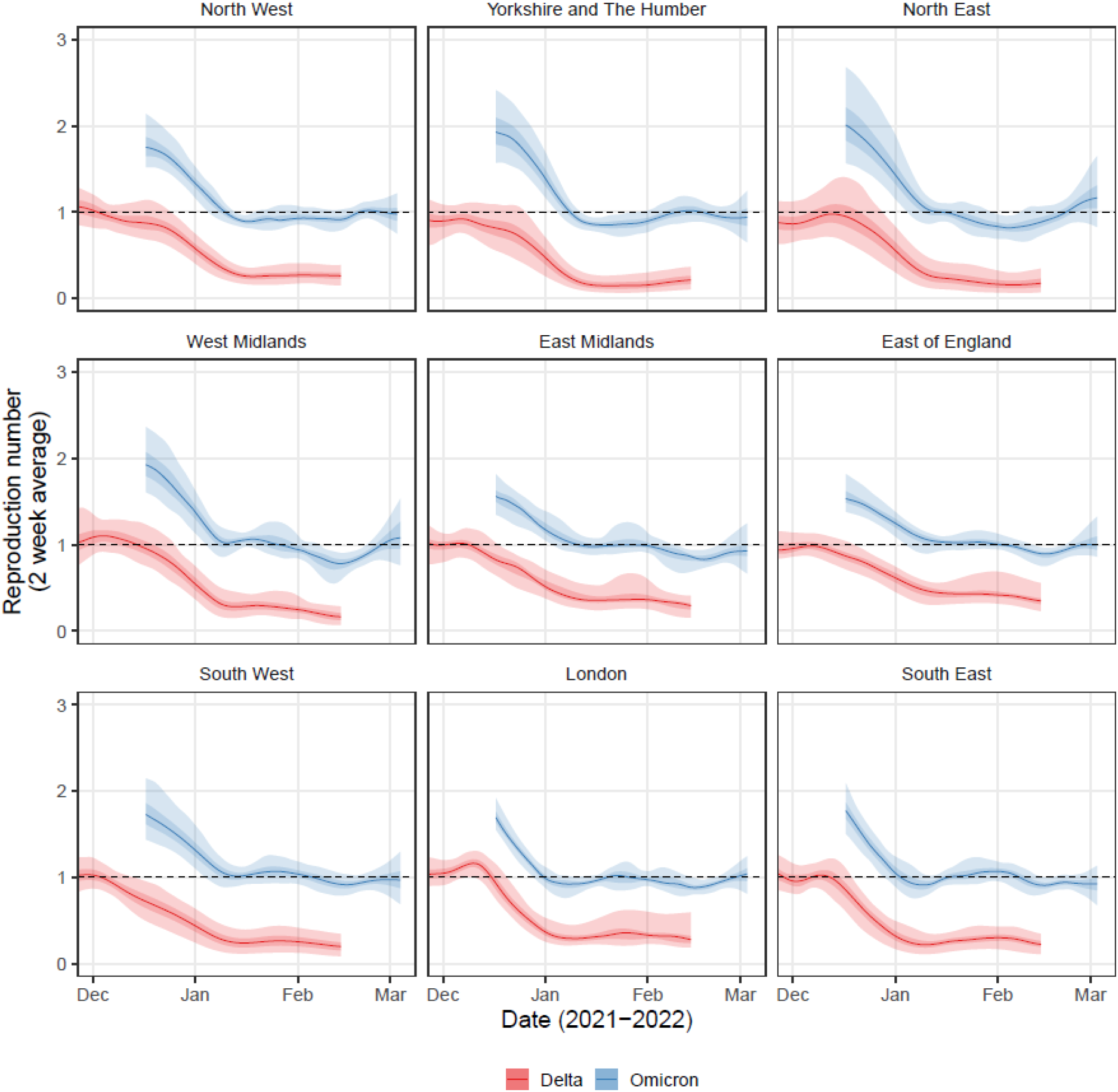
Omicron vs Delta Rt by region. Rolling two-week average (prior two weeks) Reproduction number for Omicron (blue) and Delta (red) in each region of England as inferred from mixed-effects Bayesian P-spline models. Estimates are shown with a central estimate (solid line) and 50% (dark shaded region) and 95% (light shaded region) credible intervals. Dashed line shows R=1 the threshold for epidemic growth. Estimates for each lineage are only displayed for the period over which the lineage was detected in REACT-1 samples.

**Supplementary Figure 11:**
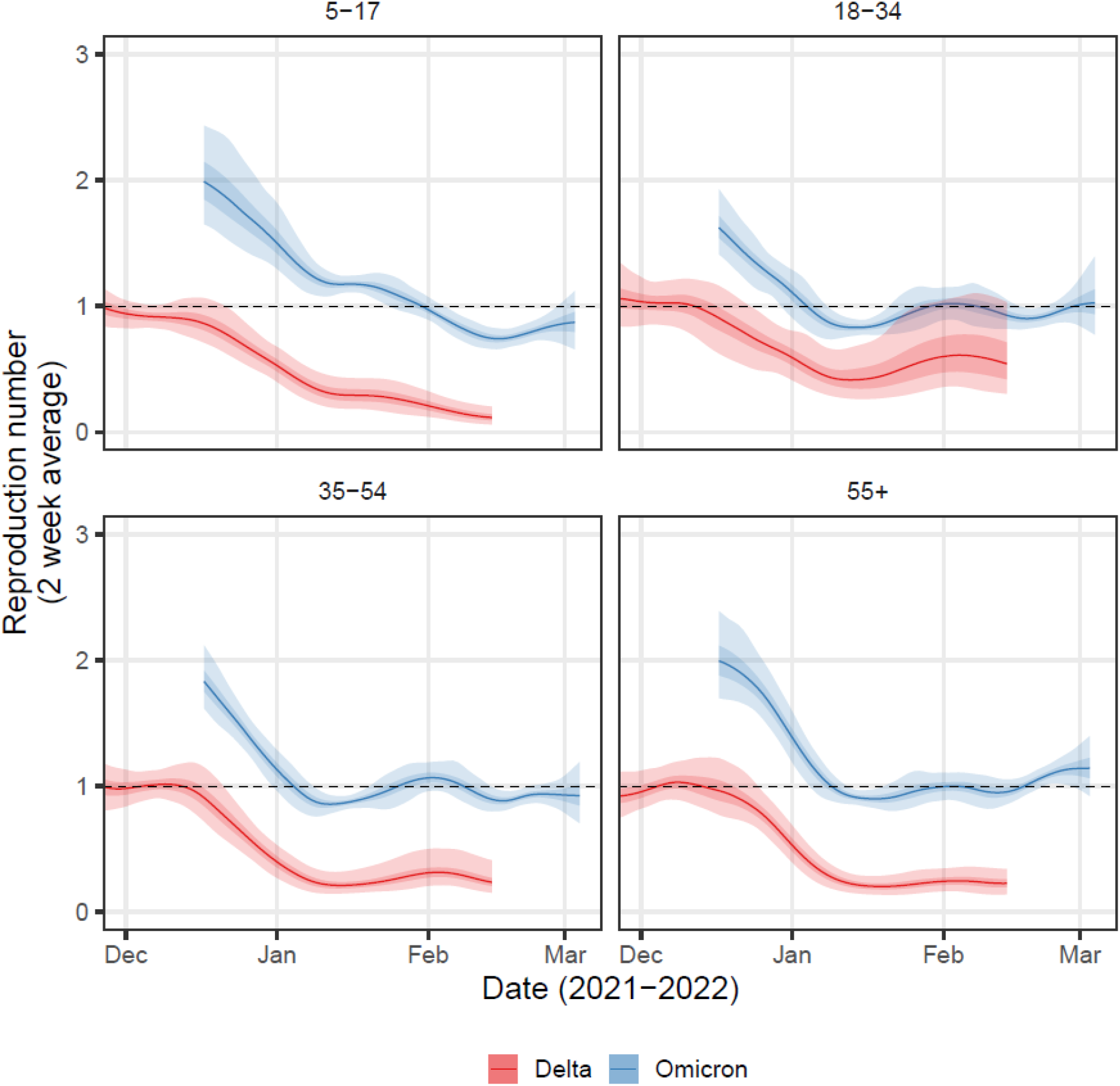
Omicron vs Delta Rt by age-group. Rolling two-week average (prior two weeks) Reproduction number for Omicron (blue) and Delta (red) for each age-group in England as inferred from mixed-effects Bayesian P-spline models. Estimates are shown with a central estimate (solid line) and 50% (dark shaded region) and 95% (light shaded region) credible intervals. Dashed line shows R=1 the threshold for epidemic growth. Estimates for each lineage are only displayed for the period over which the lineage was detected in REACT-1 samples.

**Supplementary Figure 12:**
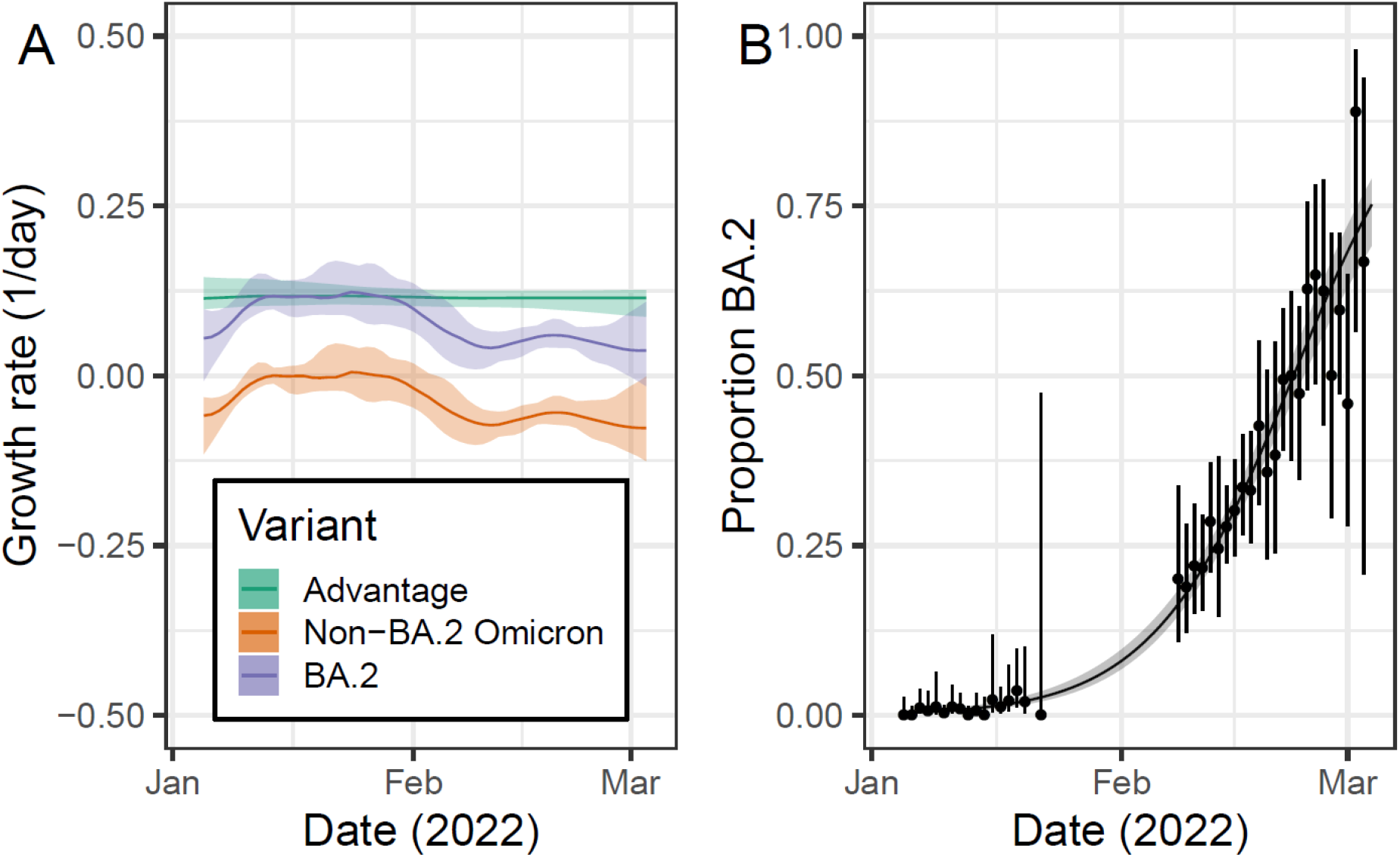
BA.2 vs non-BA.2 National growth rate + proportion graph. A) Daily growth rate of BA.2 (purple), non-BA.2 Omicron (orange) and their additive difference (green) estimated from the mixed-effects Bayesian P-spline model fitted to rounds 17 and 18 of the data. Estimates are shown with a central estimate (solid line) and 95% credible intervals (shaded region). B) Modelled proportion of lineages identified as BA.2 in England estimated using a mixed-effects Bayesian P-spline model. Estimates are shown with a central estimate (solid line) and 95% credible intervals (shaded region). Daily estimates of the proportion of lineages BA.2 (points) are shown with 95% confidence intervals (error bars).

**Supplementary Figure 13:**
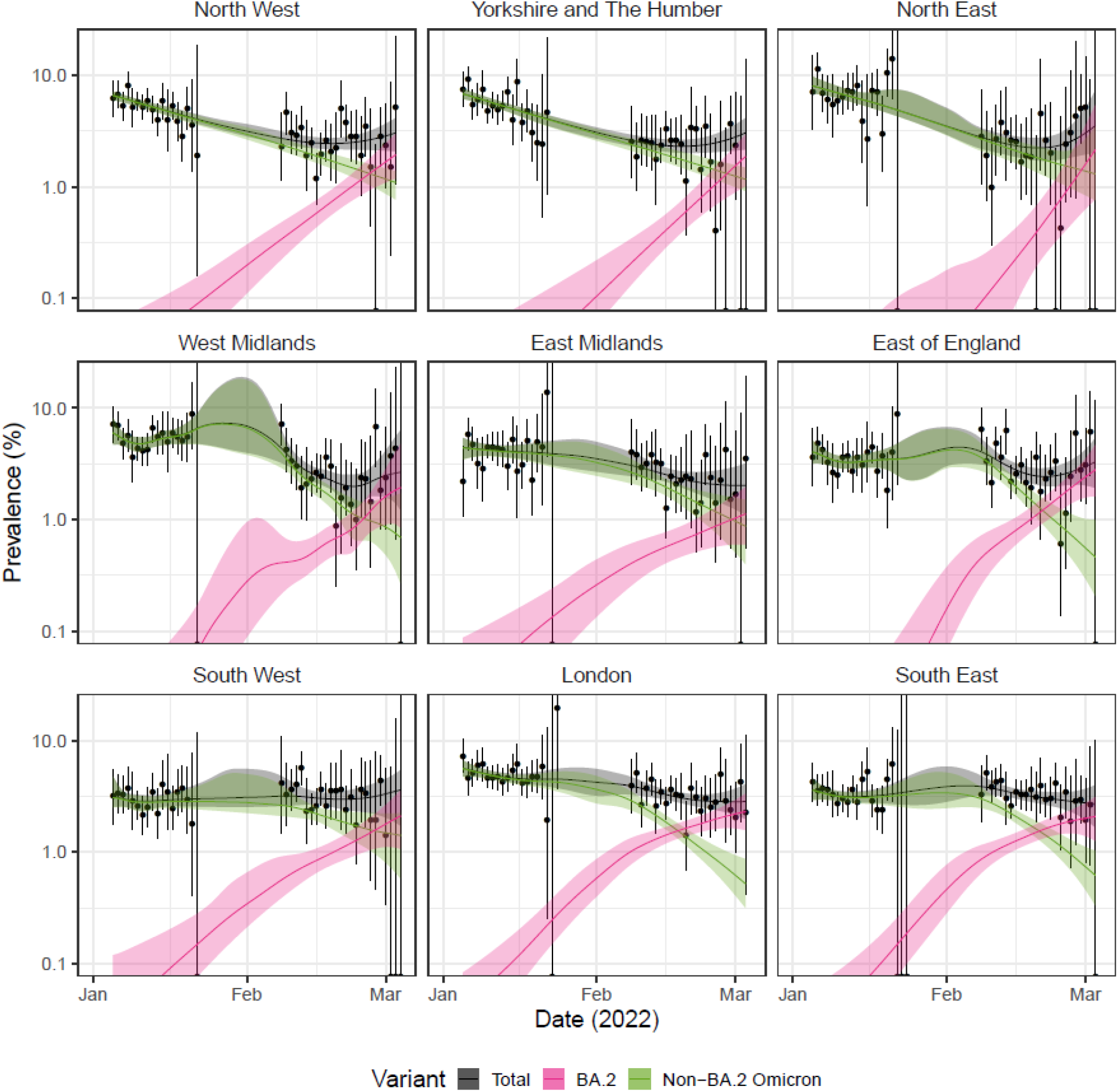
BA.2 vs non-BA.2 prevalence by region. Modelled prevalence of BA.2 (pink), non-BA.2 Omicron (green) and total prevalence (grey) in each region of England for rounds 17 and 18 estimated using mixed-effects Bayesian P-spline models. Estimates of prevalence are shown with a central estimate (solid line) and 95% (shaded region) credible intervals. Daily weighted estimates of prevalence (points) are shown with 95% credible intervals (error bars).

**Supplementary Figure 14:**
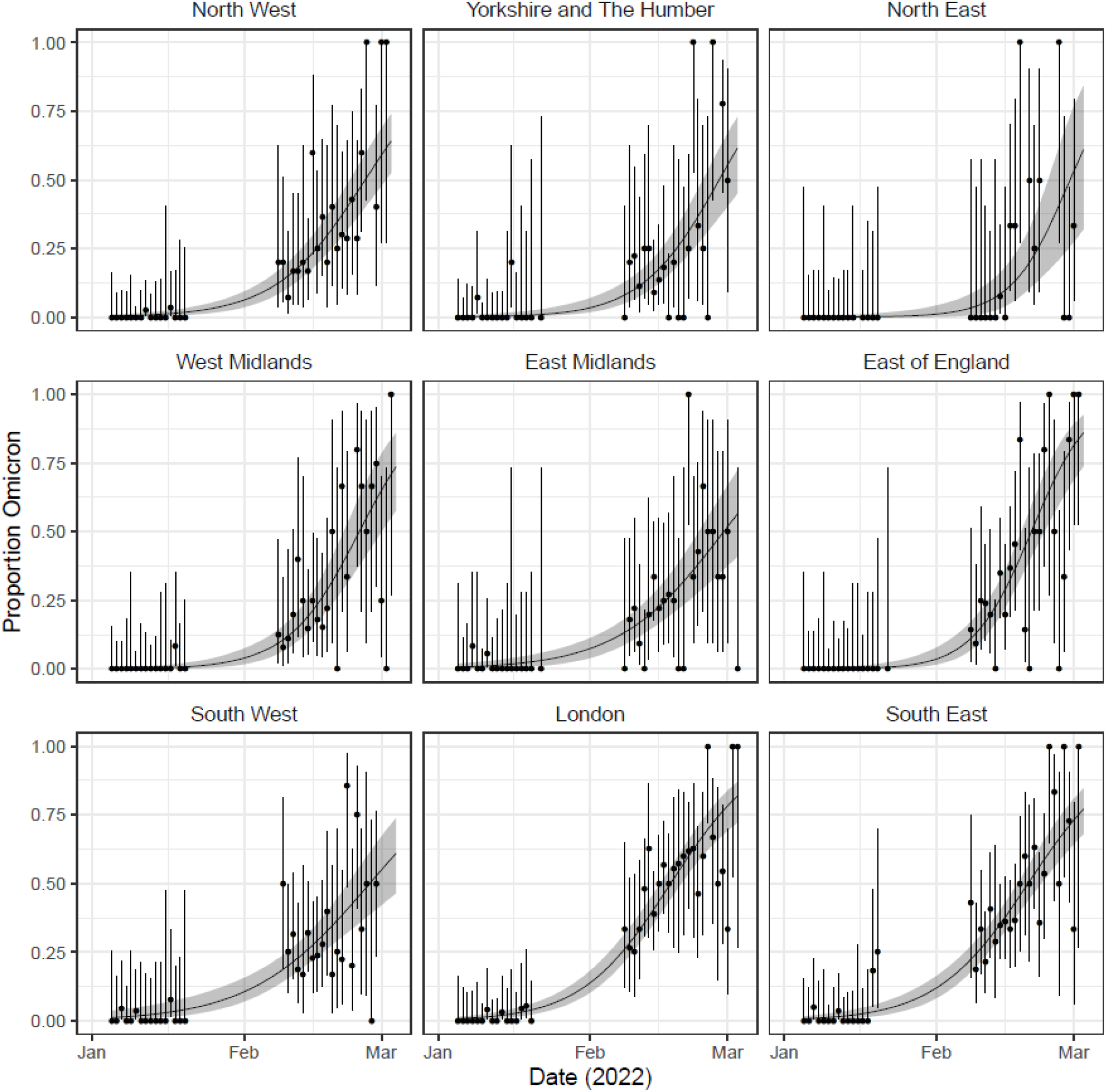
BA.2 vs non-BA.2 proportion by region. Modelled proportion of lineages identified as BA.2 by region of England for rounds 17 and 18 estimated using mixed-effects Bayesian P-spline models. Estimates are shown with a central estimate (solid line) and 95% credible intervals (shaded region). Daily estimates of the proportion of lineages BA.2 (points) are shown with 95% confidence intervals (error bars).

**Supplementary Figure 15:**
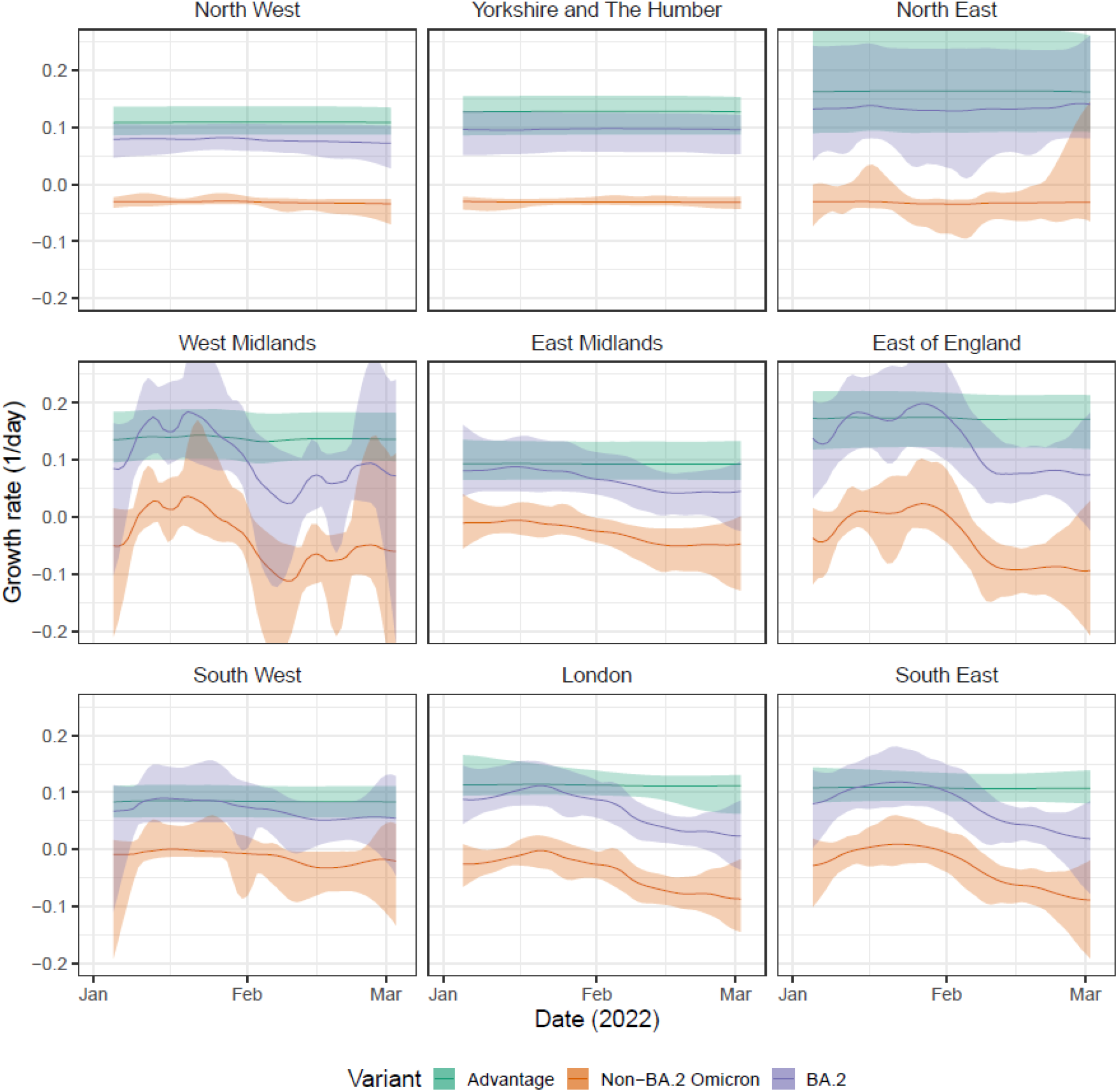
BA.2 vs non-BA.2 growth rate by region. Daily growth rate of BA.2 (purple), non-BA.2 Omicron (orange) and their additive difference (green) estimated from mixed-effects Bayesian P-spline models fitted for each region of England to rounds 17 and 18 of the data. Estimates are shown with a central estimate (solid line) and 95% credible intervals (shaded region).

**Supplementary Figure 16:**
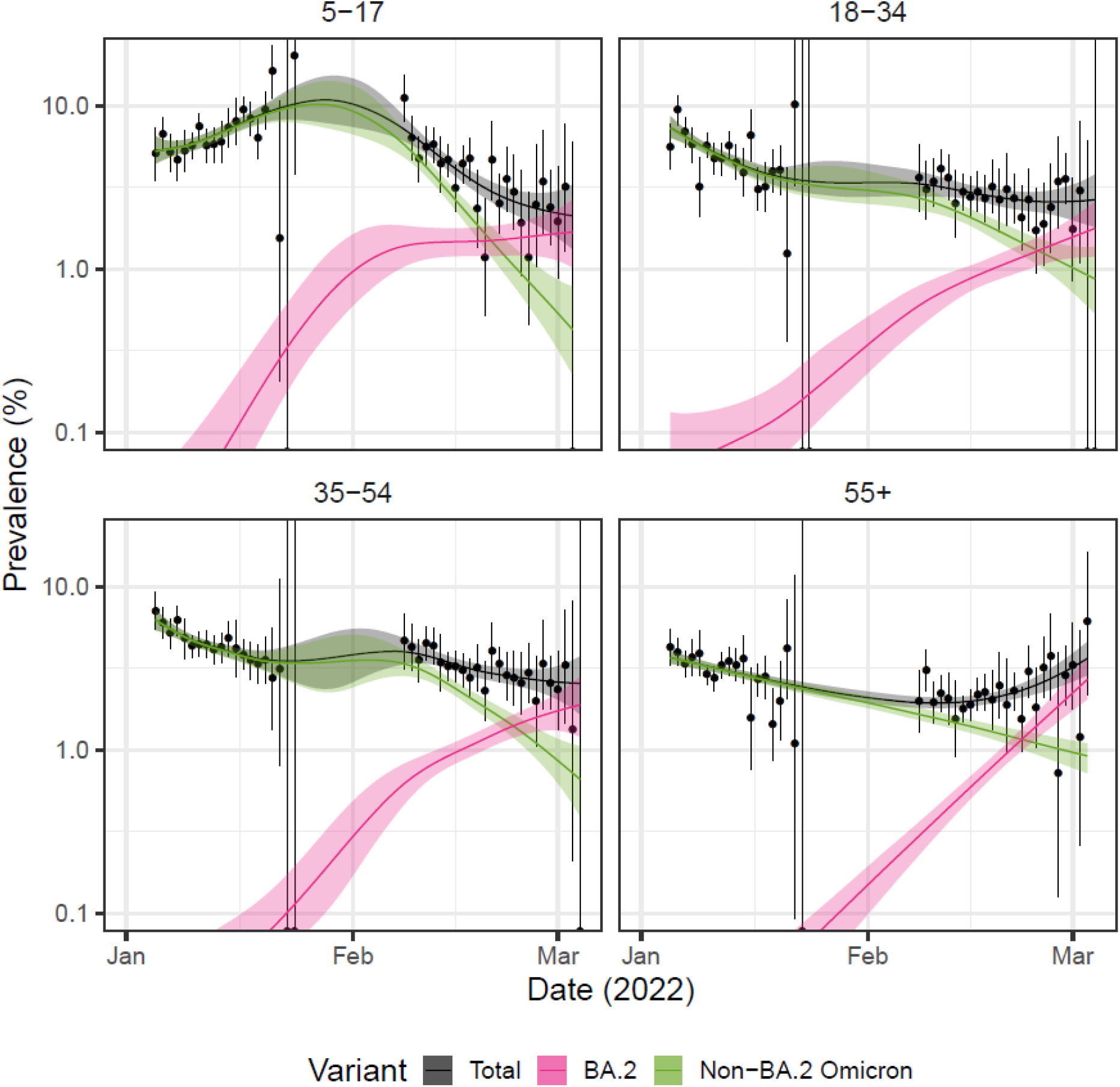
BA.2 vs non-BA.2 prevalence by age-group. Modelled prevalence of BA.2 (pink), non-BA.2 Omicron (green) and total prevalence (grey) for each age-group in England for rounds 17 and 18 estimated using mixed-effects Bayesian P-spline models. Estimates of prevalence are shown with a central estimate (solid line) and 95% (shaded region) credible intervals. Daily weighted estimates of prevalence (points) are shown with 95% credible intervals (error bars).

**Supplementary Figure 17:**
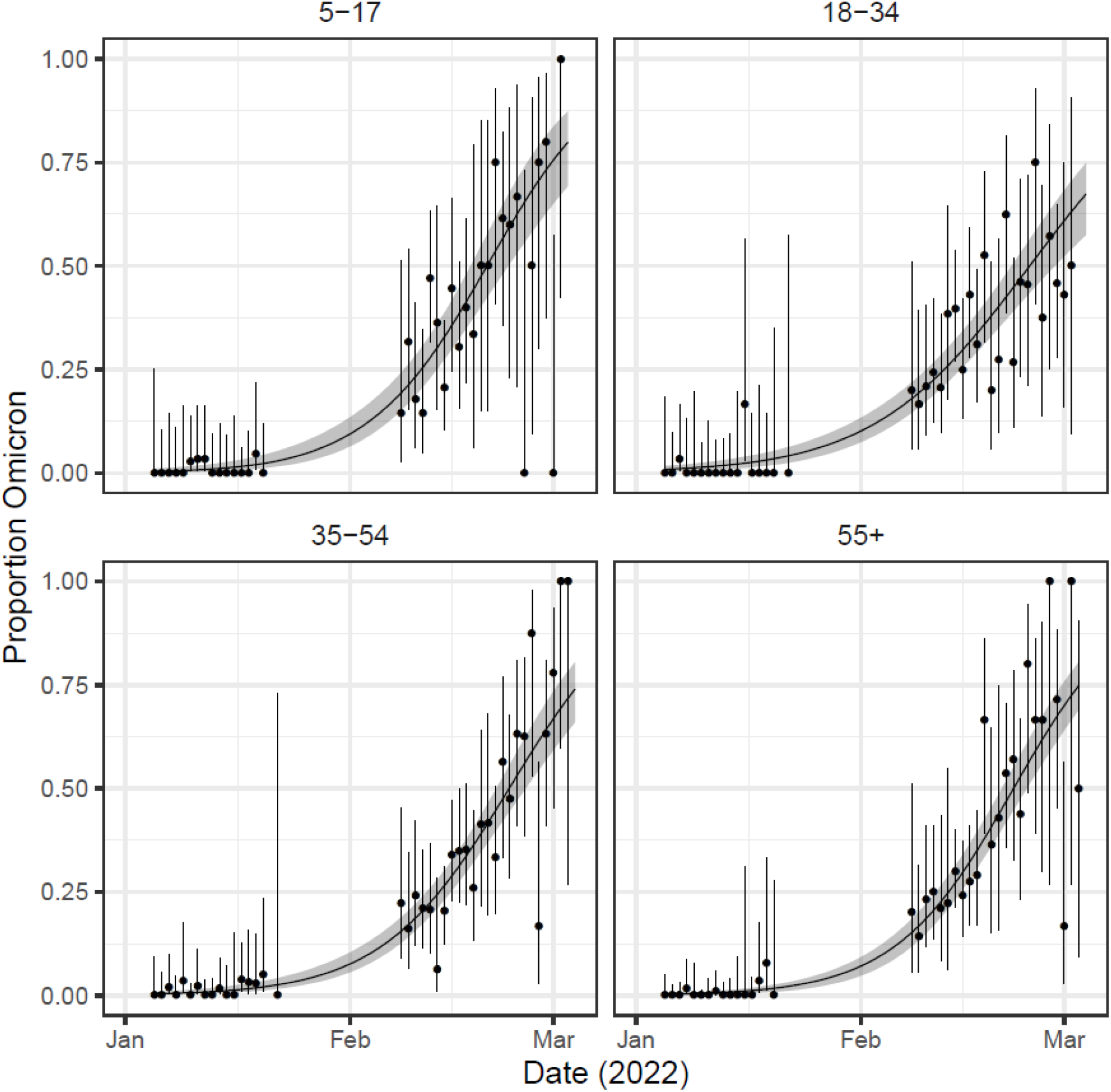
BA.2 vs non-BA.2 proportion by age-group. Modelled proportion of lineages identified as BA.2 by age-groups in England for rounds 17 and 18 estimated using mixed-effects Bayesian P-spline models. Estimates are shown with a central estimate (solid line) and 95% credible intervals (shaded region). Daily estimates of the proportion of lineages BA.2 (points) are shown with 95% confidence intervals (error bars).

**Supplementary Figure 18:**
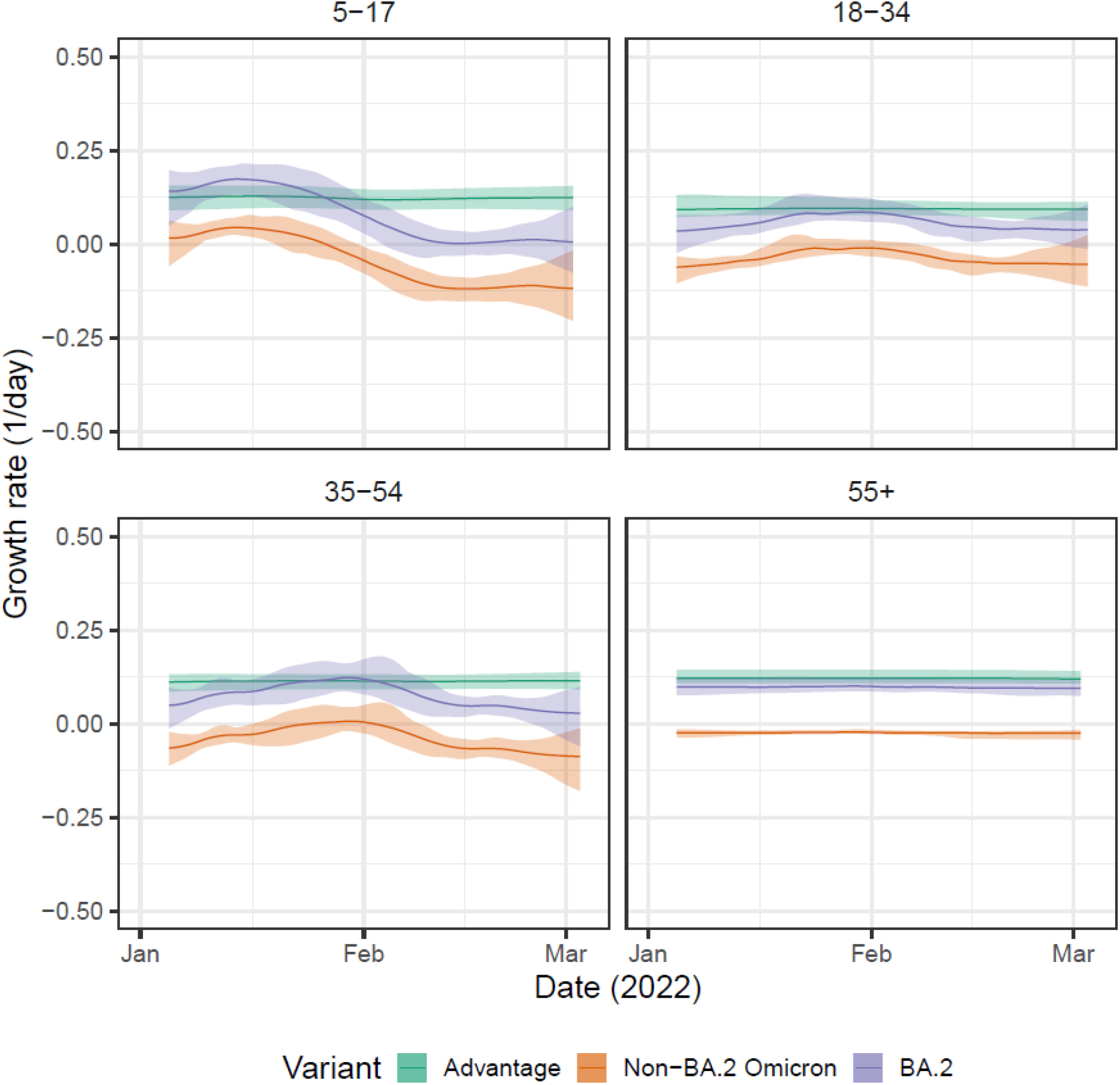
BA.2 vs non-BA.2 growth rate by age-group. Daily growth rate of BA.2 (purple), non-BA.2 Omicron (orange) and their additive difference (green) estimated from mixed-effects Bayesian P-spline models fitted by age-group in England to rounds 17 and 18 of the data. Estimates are shown with a central estimate (solid line) and 95% credible intervals (shaded region).

**Supplementary Figure 19:**
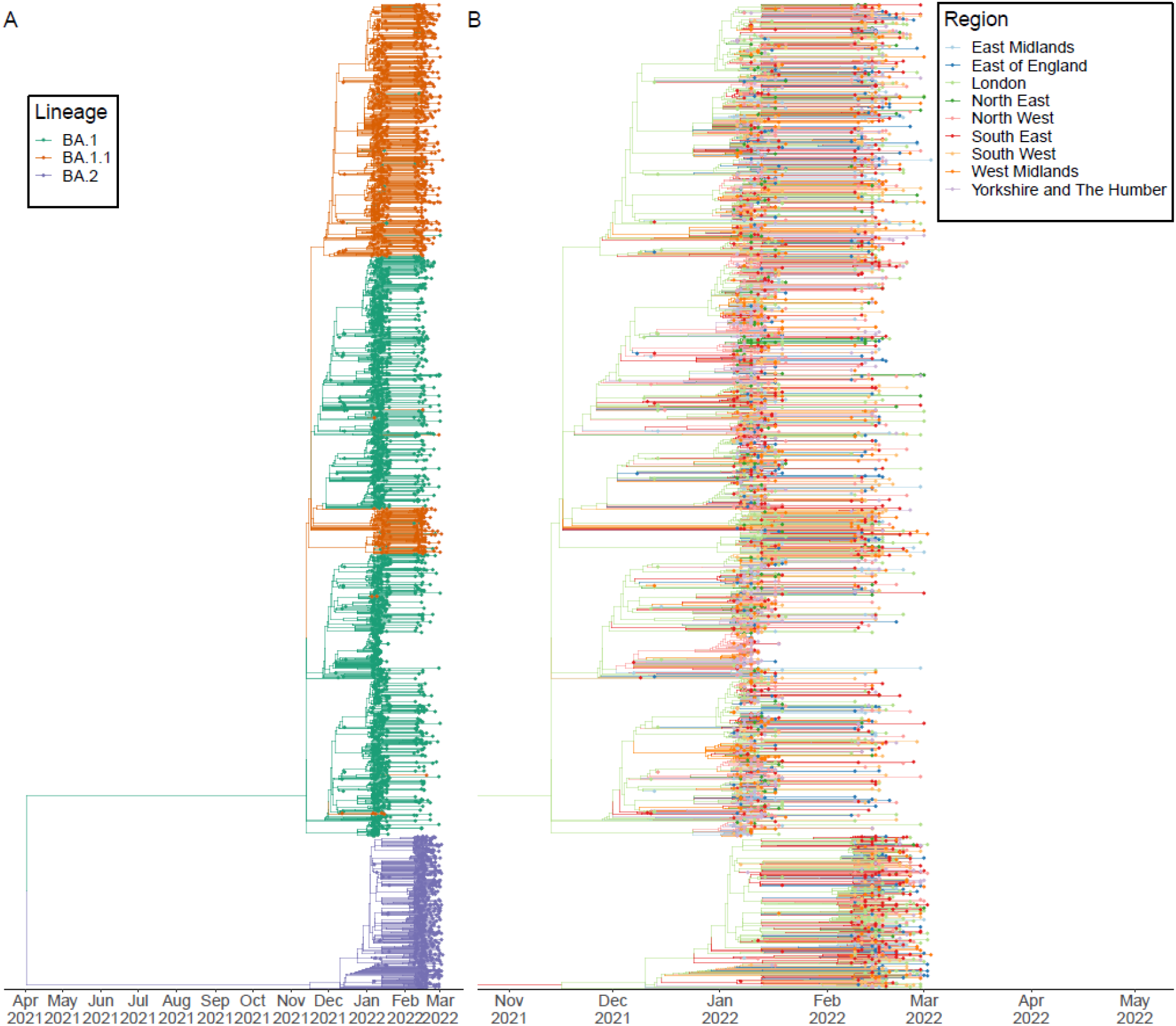
Phylogeographic model of Omicron. (A) Time-resolved phylogenetic tree for all Omicron lineages. Tips and nodes have been coloured by their inferred Omicron sub-lineage. (B) Time-resolved phylogenetic tree for all Omicron lineages. Tips have been coloured by the region in which they were collected. Nodes have been coloured by their inferred region (using a mugenic model). Note we have only shown the tree from November 2021 onwards.

**Supplementary Figure 20:**
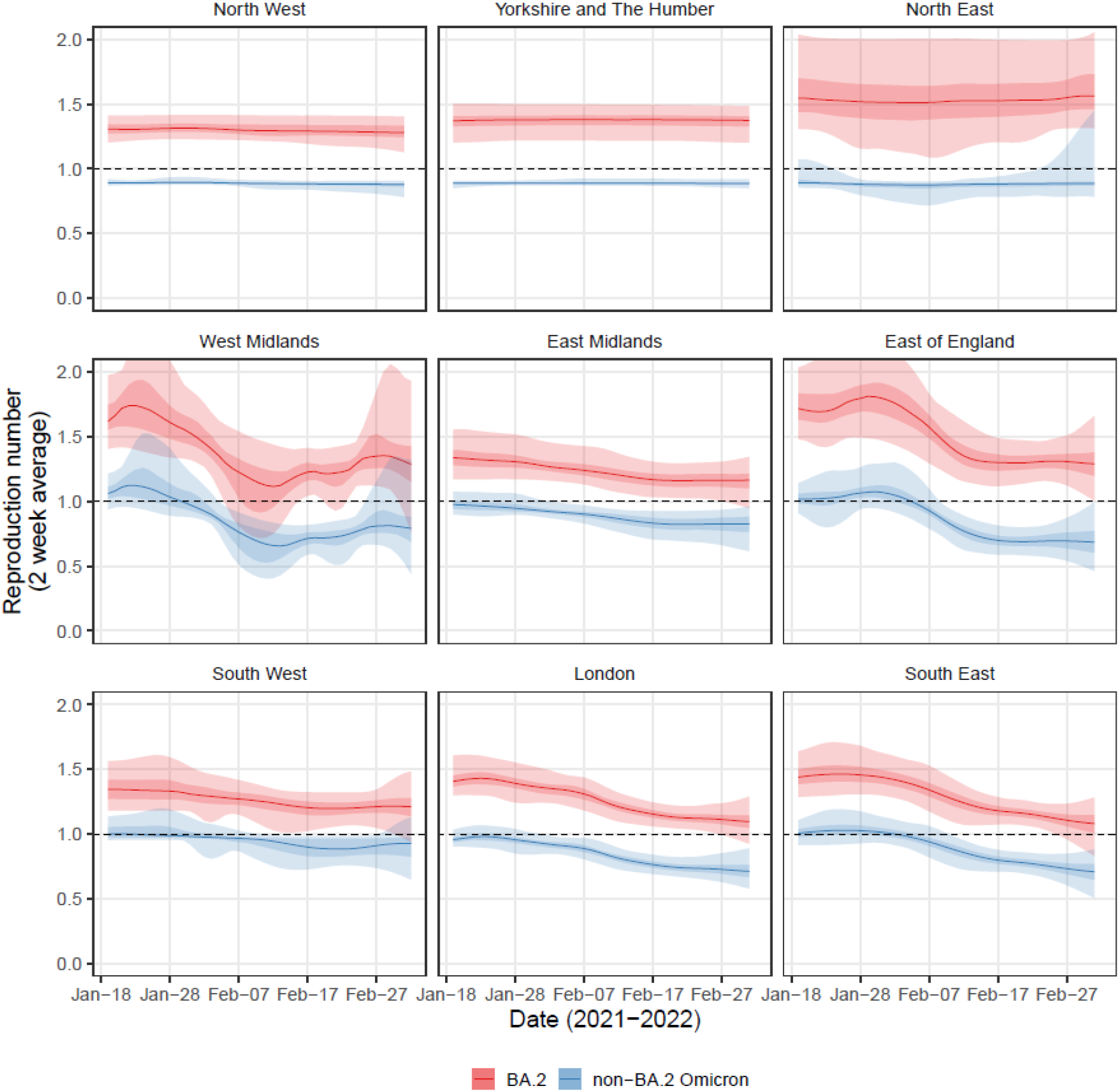
BA.2 vs non-BA.2 Rt by region. Rolling two-week average (prior two weeks) Reproduction number for BA.2 (red) and non-BA.2 Omicron (blue) in each region of England as inferred from mixed-effects Bayesian P-spline models fitted to rounds 17 and 18 of the data. Estimates are shown with a central estimate (solid line) and 50% (dark shaded region) and 95% (light shaded region) credible intervals. Dashed line shows R=1 the threshold for epidemic growth.

**Supplementary Figure 21:**
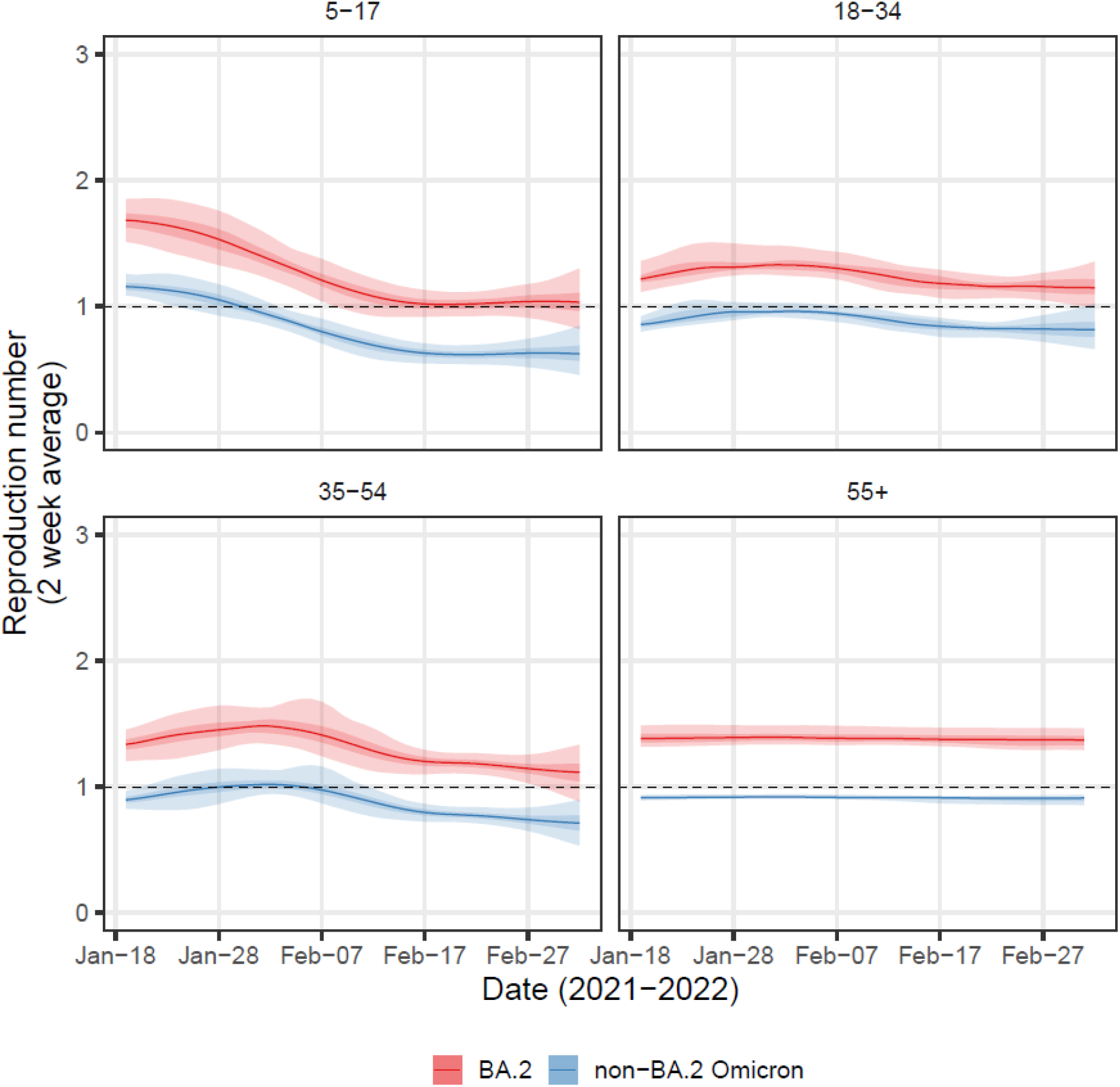
BA.2 vs non-BA.2 Rt by age-group. Rolling two-week average (prior two weeks) Reproduction number for BA.2 (red) and non-BA.2 Omicron (blue) for each age-group in England as inferred from mixed-effects Bayesian P-spline models fitted to rounds 17 and 18 of the data. Estimates are shown with a central estimate (solid line) and 50% (dark shaded region) and 95% (light shaded region) credible intervals. Dashed line shows R=1 the threshold for epidemic growth.

## Notes

### Competing Interest Statement

The authors have declared no competing interest.

